# Minocycline attenuates panicogenic responses in a CO_₂_-induced panic attack model: a translational approach

**DOI:** 10.64898/2025.12.05.25341708

**Authors:** Beatriz F. G. de Oliveira, Laiana A. Quagliato, Alana T. Frias, Luis Gustavo A. Patrone, Elisa M. Fonseca, Maria Emanuelle Reis, Breno Vilas Boas Raimundo, Karen Cristina Oliveira, Mariana Marchi Santoni Biasioli, Caroline Maria Marcos, Tatiana Maria de Souza-Moreira, Felipe Dalvi-Garcia, Natia Horato, Kênia C. Bícego, Hélio Zangrossi, Alexandra Ivo de Medeiros, Antonio E. Nardi, Luciane H. Gargaglioni

**Affiliations:** Dept of Animal Morphology and Physiology, School of Agricultural and Veterinary Sciences, São Paulo State University (UNESP), Jaboticabal, SP, Brazil; Laboratory of Panic and Respiration (LABPR); Institute of Psychiatry of Federal University of Rio de Janeiro (UFRJ). Rio de Janeiro, RJ, Brazil; Department of Biological Sciences. School of Pharmaceutical Sciences. São Paulo State University (UNESP), Araraquara, SP, Brazil; Laboratory of Biology, Control and Surveillance of Insect Vectors, Oswaldo Cruz Institute, FIOCRUZ/RJ, Rio de Janeiro, Brazil; Department of Pharmacology, School of Medicine of Ribeirão Preto, University of São Paulo, Ribeirão Preto, SP, Brazil

**Keywords:** hypercapnia, panic disorder, microglia, clonazepam, cytokines

## Abstract

There is a connection between neuroinflammation and panic attacks (PA), as microglia-driven pro-inflammatory responses help detect homeostatic disturbances like CO_₂_ inhalation. This model has become widely used in research since CO_₂_ exposure can trigger PA in humans and panic-related behavior in mice. Minocycline inhibits microglia activation, serving as a promising tool to attenuate CO_2_-induced PA. The locus coeruleus (LC) is a CO_₂_/pH-sensitive region, and disruptions in its activity are linked to psychiatric conditions such as panic disorder (PD). We investigated the involvement of microglia in the respiratory and behavioral responses induced by CO_2_ in mice and the effect of minocycline and clonazepam treatment. We also assessed in mice whether LC microglia are activated after hypercapnia using IBA-1 immunohistochemistry. Translationally, PD patients were treated with minocycline and clonazepam and examined for their CO_2_-responsiveness. LC microglia were activated 6 hours after exposure to 20% CO_2_ in mice. This panicogenic stimulus also induced hyperventilation as well as active panic-related escape responses, characterized by jumps and running episodes. Minocycline and clonazepam decreased escape expression during the CO_2_ challenge, but only the former drug reduced hyperventilatory responses. None of the drugs changed IL levels in LC. In humans, minocycline reduced the severity of CO_2_-induced panic attacks and also modulated the immune response by lowering IL-2sRα and increasing IL-10 levels. Exposure to hypercapnia activates microglia in the LC of mice. Treatment with minocycline, similar to the clinically effective panicolytic clonazepam, attenuates CO_2_-induced panic-like responses in both mice and humans. These results support the potential of minocycline as a therapeutic strategy for PD.

## 1. Introduction

Over the years, an extensive line of research has been structured to understand the connection between respiratory disturbances and anxiety disorders, including panic attacks (1,2). Panic disorder (PD) represents a mentally and physically debilitating type of anxiety disorder characterized by unexpected and recurrent panic attacks (PAs), often accompanied by intense fear and associated with cardio-respiratory symptoms (DSM-5). The neurobiological pathways underlying panic attacks have been the subject of many studies (3–5), and there is evidence to suggest that respiratory disturbances play a critical role in their occurrence (6,7).

PAs and anxiety have been evoked in patients with panic disorder who underwent respiratory challenge tests (8–10). Over the years, these respiratory tests, such as CO_2_ inhalation, have become an efficient tool for deepening the studies of the respiratory subtype of panic attack, with the hypercapnic environment being a reliable panicogenic agent (11–15). We know that as CO_2_ concentrations increase, the threat of asphyxia or suffocation becomes more imminent, triggering defensive responses. In mice, for example, inhalation of high concentrations of CO_2_ induces panic-related escape responses, which are reduced by standard panicolytic drugs such as alprazolam and chronic fluoxetine (15).

The respiratory mechanisms underlying CO_2_-evoked PAs involve a strong participation in the chemosensory responses of the central nervous system. Changes in O_2_ and/or CO_2_ concentrations will trigger rapid ventilatory adjustments so that arterial blood gases return to their set point. One of the central chemoreceptive structures is the locus coeruleus (LC), a region located in the brainstem responsible for releasing noradrenaline to various structures, as well as participating in fundamental functions such as respiratory control through its CO_2_ chemosensitivity (16,17). Some patients with PD exhibit altered noradrenergic function (18), and studies have shown that the reduction and/or increase in CO_2_ concentrations altered the firing rate of these neurons by up to 53% (19,20). In humans, some studies have shown that pupil dilation due to stress situations is considered a non-invasive marker of LC noradrenergic activity (21).

In addition to neuronal involvement, glial cells are also implicated in the mechanisms underlying anxiety disorders, and some studies suggest that changes in microglial-driven processes play a role in the pathophysiology of different neurological and psychiatric disorders (22,23). Various disturbances to homeostasis—such as inflammation, neurodegenerative diseases, or traumatic injury—can trigger dynamic structural responses in microglia, characterized by alterations in cell body size, retraction or extension of processes, and changes in motility. These morphological alterations, which range from a ramified “surveying” state to an amoeboid “activated” phenotype, are often mediated by signaling pathways involving ATP, cytokines, and pattern recognition receptors such as Toll-like receptors (TLRs) (24,25). Such alterations enable microglia to shift from homeostatic surveillance to roles in phagocytosis, antigen presentation, and cytokine secretion (24,25), detecting an imbalance, such as in ionic homeostasis, for example (26,27).

Minocycline, a second-generation derivative of tetracycline with bacteriostatic antimicrobial action, is also known to have anti-inflammatory properties that are independent of its antibacterial action, exerting a neuroprotective function, limiting inflammation and oxidative stress (28). It has been shown that administration of minocycline in mouse models of amyotrophic lateral sclerosis attenuated the induction of expression of inflammatory microglia markers (pro-inflammatory state) (29). At the same time, it did not affect the transient increase in the expression of anti-inflammatory microglia markers (anti-inflammatory) during the early phase of the disease (29). In a more recent study, Rooney et al. (30) reported that minocycline attenuated anxiety-like behaviors in mice. This attenuation occurred through modulation of microglial activity within the dentate gyrus. The authors suggest that medications with anti-inflammatory properties targeted at microglia may hold promise as potential complementary anxiolytic alternatives for individuals predisposed to anxiety disorders.

Here, we first addressed whether exposure to a severe hypercapnic challenge modifies the microglial phenotype in the LC of mice. As microglia may play a significant role in the genesis/regulation of anxiety and possibly panic attacks, we next investigated the involvement of these cells, through minocycline, in adult mice’s respiratory and behavioral responses during exposure to CO_2_. Finally, we investigated the effects of this drug on anxiety and panic responses of PD patients exposed to the same respiratory challenge. In mice and humans, we compared the effects of minocycline to those caused by a standard anxiolytic drug, clonazepam.

## 2. Methods and Materials

### 2.1 Mice studies

#### 2.1.1 Animals and minocycline and clonazepam treatment

Male and female C57BL/6 mice obtained from the University of Sao Paulo, Campus of Ribeirao Preto, 8–12 weeks old on the day of the experiment, were housed in groups of 5 per cage maintained under standard laboratory conditions (25 ± 1 °C and 12:12 h light: dark cycle, lights on at 06:30), with food and water available ad libitum. The animals were randomly divided into groups. All experiments were conducted with the approval of the local College of Agricultural and Veterinary Sciences Animal Care and Use Committee (CEUA-FCAV-UNESP-Jaboticabal; Protocol: n°11.794).

Minocycline (Hovione Farmaciencia, Portugal) and clonazepam (Roche, Brazil) were dissolved in 0.1 M PBS and prepared freshly before injection.

#### 2.1.2 Microglia analyses

Microglial morphological analyses have been previously used in studies for evaluating the activity of these cells, through cell density, morphological index (cell body area and branching area), and nearest neighbor distance (NND) (Figure 1A).

**Figure 1:**
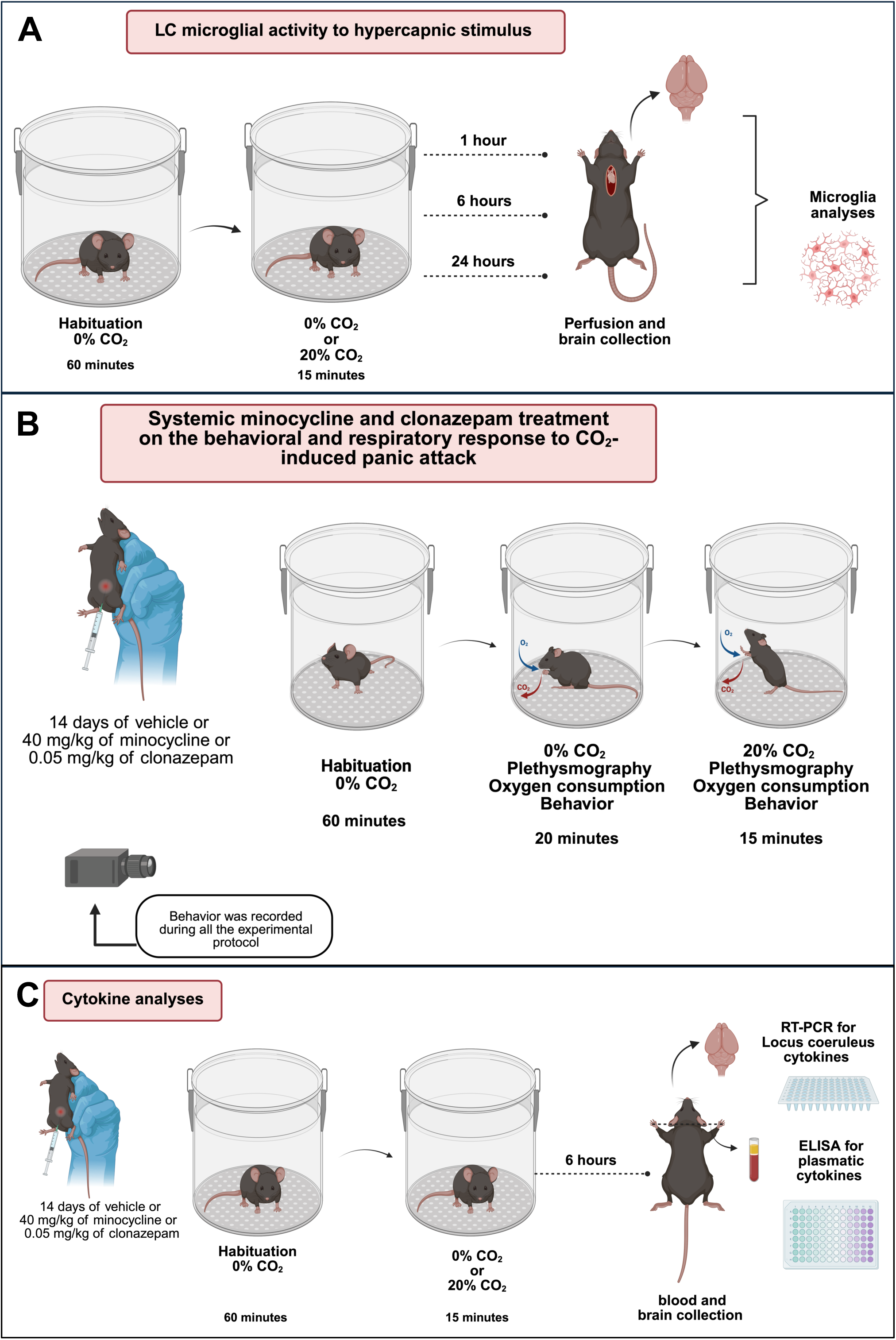
Schematic illustration of the experimental protocols for. (**A**) microglial evaluation following CO_₂_ exposure; (**B**) respiratory and behavioral recordings, and (**C**) cytokine analysis, both performed after treatment with minocycline, clonazepam, or vehicle under normocapnic and hypercapnic conditions.

For the analysis of microglial cell density in the LC, we used a 20X magnification objective, guided by the mouse brain atlas (31), along with immunohistochemistry for tyrosine hydroxylase (TH), to delimit the LC region (Figure 2). This procedure facilitated the construction of a representative drawing outlining the LC region, which was used in all photomicrographs labeled with the Iba-1 antibody.

**Figure 2:**
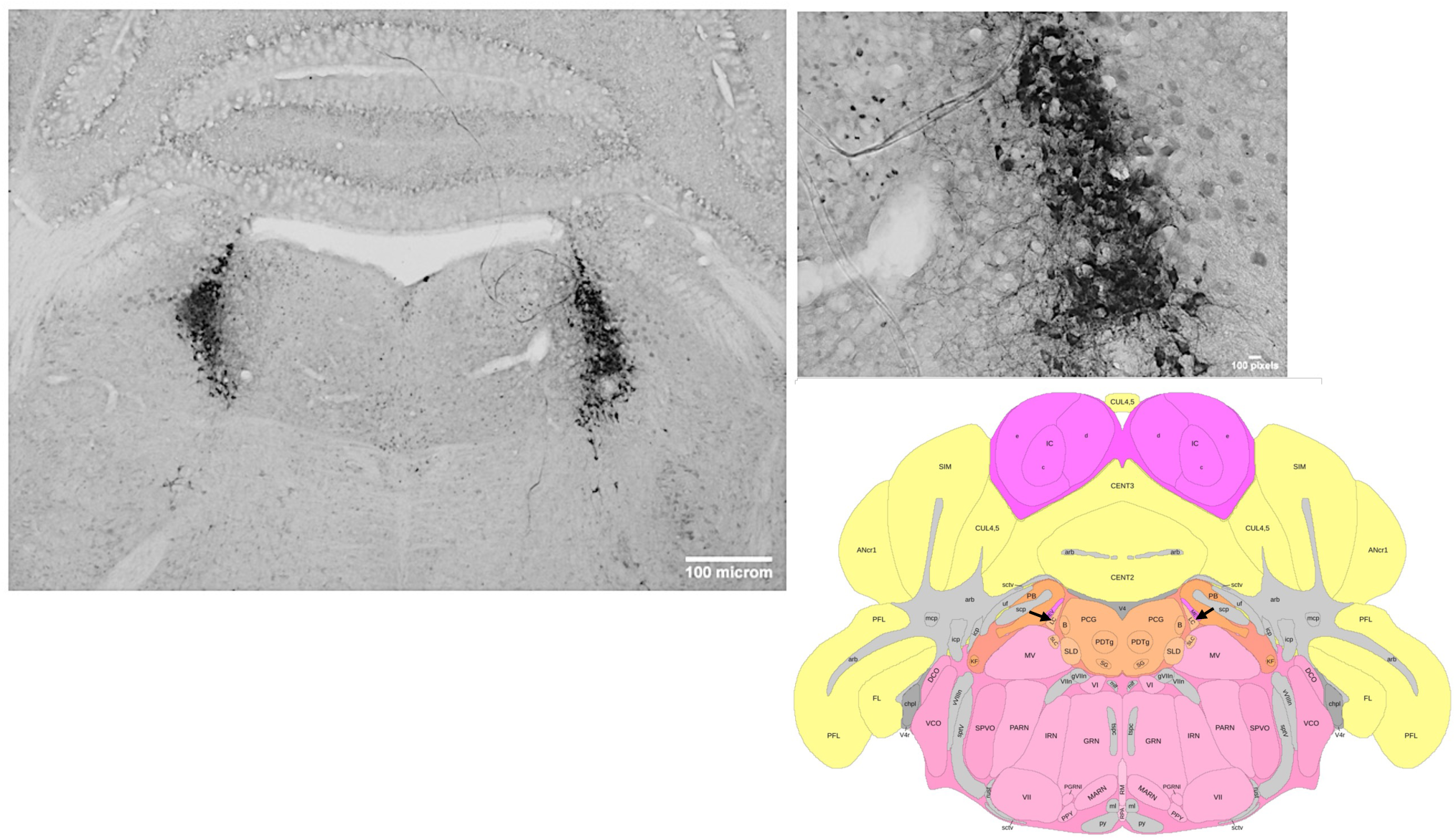
Photomicrograph of immunohistochemistry for tyrosine hydroxylase (TH) to delimit the Locus coeruleus (LC) region in adult mice. Objective of 10x (**A**), 20x (**B**), and schematic drawing of the LC location, adapted from Allen Brain Cell Atlas (**C**). Scale bar: 100 μm.

For brain tissue collection, the animals were deeply anesthetized with isoflurane and perfused through the left ventricle with the aid of a peristaltic pump (Masterflex; Cole-Parmer Instrument, USA) with phosphate-buffered saline (PBS: 0.01 M, pH 7.4) followed by 4% paraformaldehyde (PFA) solution diluted in phosphate buffer (PB: 0.2 M). The brains were removed, fixed for 24 hours in 4% PFA at 4°C, placed in 30% sucrose for 48 hours at 4°C, frozen in isopentane, and stored in a freezer at –20°C. The brains were sectioned using a cryostat (model CM1860, Leica, Germany) into 40-μm coronal sections that contained the LC region. The sections underwent an immunohistochemistry protocol for labeling ionized calcium-binding adapter molecule 1 (Iba-1, anti-IBA I antibody, Fujifilm Irvine Scientific, USA, catalog number: 019-19741), a selective marker for microglia, as previously described (32–34).

The number of microglial cells in the region was quantified using ImageJ software. The cell density was obtained by the ratio of the total number of cells in the analyzed area (35, 36). Both the left and right sides of the LC were analyzed for all parameters. In the ImageJ software, a drawing was manually created around the analyzed cell, outlining both the arborizations and cell bodies, using a 40X magnification objective. This analysis process was repeated for at least the 10 nearest cells, calculating each cell’s cell body area and arborization area. From the averages of these variables and the ratio between the cell body area and arborization area, the morphological index was determined. This parameter indicates morphological changes in microglia (36), allowing us to determine whether the considered variables in the analysis favor the “hypertrophic” (activated) form of microglial cells compared to the ramified (or “surveillant”) form (36,37).

Following the cellular morphology definition, all analyzed cells and those nearby were selected for evaluation using the ‘NND’ (nearest neighbour distance) tool within the ImageJ software. This parameter measures the distance from each microglial cell to its nearest neighbor, indicating cell distribution and motility (38).

#### 2.1.3. Panic attack induction and behavioral analyses

The 20% CO_2_ air challenge was used to induce panic behavioral and ventilatory responses in mice (15; 39). To promote a hypercapnic environment, a gas mixer (Gas mixer GSM-3, CWE Inc., USA) was used to produce the following gas mixture: 21% O_2_, 20% CO_2_, and 59% N_2_. The CO_2_ concentration was gradually increased over 15 minutes until it reached a level of 20%.

Two cameras (Logitech, USA) recorded the experiment laterally and above the experimental chamber. The behavioral responses during CO_2_ exposure were measured offline by a trained researcher who was blind to the drug treatment. The number of jumps towards the ceiling of the cage or running episodes inferred the panic-like escape response. We also computed the percentage of time spent in freezing behavior (total lack of movement except for breathing). Locomotion was measured automatically by a video tracking system (AnyMaze, version 4.96, USA) for 20 minutes, which preceded exposure to CO_2_, with the animals breathing room air.

#### 2.1.4 Respiratory measurements and O_2_ consumption

Ventilation (V_E_) in mice was measured by whole-body plethysmography (39). The animal was placed in an experimental chamber (5 L) for a minimum habituation of 60 minutes with normocapnic air. A flow meter model (“Mass Flow System” – MFS, Sable Systems International, Inc., Las Vegas, USA) maintained the airflow at approximately 1.8 L/min. After the end of the habituation period, two ventilatory measurements were taken under room air conditions, with a 10-minute interval between each measurement. Afterward, the animal was exposed to hypercapnic air (20% CO_2_, 21% O_2_, N_2_ balance) for 15 minutes, and, in the end, a new measurement of V_E_ was obtained (Figure 1B). The oxygen consumption (VO_2_) was measured using the indirect calorimetry method (push mode configuration), an open respirometry system, to obtain metabolic inferences. For the VO_2_ measurements, the air in the chamber was sampled throughout the experiment, except during the V_E_ recordings, when the chamber was sealed (Figure 1B).

Using a transducer to measure the pressure differential (TSD 160A, Biopac Systems, Santa Barbara, CA – USA), the signals were collected and passed to an amplifier (DA 100C, Biopac Systems, Inc., USA). Further, these signals were directed to an analog-to-digital converter and digitized on a computer equipped with the AcqKnowledge acquisition software (MP 100, BioPac Systems, Inc., Santa Barbara, CA, USA). We injected the same amount of air (1 mL) into the chamber for the volume calibration using a graduated syringe for each experiment. V_E_ was calculated by multiplying respiratory frequency (f_R_) by tidal volume (V_T_). Tidal volume was calculated using the Drorbaugh and Fenn formula (40):

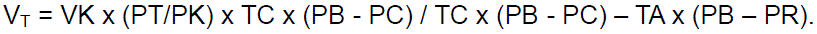

In the Drorbaugh and Fenn formula, PT represents the pressure deflection corresponding to each tidal volume (V_T_), PK corresponds to the pressure deflection associated with the injection of the calibration volume (VK), TA indicates the air temperature inside the animal chamber, and PC is the water vapor pressure in the animal chamber. At the same time, Tb is the body temperature, and PR indicates the water vapor pressure at Tb. The PC and PR were calculated using Dejours’s (41) table. V_E_ and V_T_ values were presented under standard conditions, accounting for ambient barometric pressure conditions at body temperature (Tb) and saturated with water vapor (BTPS).

The oxygen consumption (VO_2_) was measured using the indirect calorimetry method (push mode configuration), an open respirometry system, to obtain metabolic inferences. An aquarium pump was used to direct the airflow inside the chamber for room air exposure (normocabic normoxia), while the hypercapnic gas mixture (20% CO_2_, 21% O_2_, and N_2_ balance) was pumped by a gas mixer (Gas mixer GSM-3, CWE Inc., USA). The exhaled gas was dried through a column of Drierite (W. A. Hammond Drierite Co. Ltd, Xenia, OH, USA) and passed to the oxygen analyzer (model ML206, ADInstruments®, Australia). The O_2_ analyzer sampled the air during the experiments to determine VO_2_ by the Power-Lab data acquisition program (ADInstruments®/Chart Software, version 7.3, Sydney, Australia).

#### 2.1.5 Quantification of plasmatic cytokines by ELISA

Blood was collected via decapitation using a guillotine and immediately transferred into heparinized tubes (Figure 1C).

The samples were centrifuged at 10,000 × g for 10 minutes using an Eppendorf centrifuge 5403 (Germany). Plasma was stored at −80 °C in a bio ultra-freezer (Snijders Scientific, Netherlands) until further analysis. Subsequently, the Quantification of IL-6, IL-10, TNF-α, and IL-2sRα was performed using an enzyme-linked immunosorbent assay (ELISA) (BD), following the manufacturer’s instructions. The detection limits for each cytokine are specified in Table 1.

**Table 1.** Cytokines Evaluated with Their Respective Detection Limits.

| Cytokine | Detection limit | Manufacturer | Catalog |
| --- | --- | --- | --- |
| BD OptEIA™ Set Mouse IL-6 | 15.6 pg/mL | BD | #2653KI |
| BD OptEIA™ Set Mouse IL-10 | 31.2 pg/mL | BD | #2657KI |
| ELISA MAX™ Mouse TNF-α | 7.8 pg/mL | Biolegend | #430904 |
| BD OptEIA™ Set Mouse IL-2sRα | 7.8 pg/mL | BD | #559104 |

##### Cytokine expression by RT-qPCR analysis

Following decapitation, the whole brain was rapidly removed and frozen in a cryostat (model CM1860, Leica, Germany) at –22°C and coronally-sectioned to find the stereotaxic coordinates of the LC (distance from bregma: – 10.3Lmm to –9.3 mm). Samples of 1.0 mm thickness were removed using a 10-gauge needle. The LC was dissected precisely, weighed, and transferred into pre-cooled Eppendorf tubes filled with an RNA-later solution and kept at – 80°C freezer (Panasonic, model MDF-U56VC-PA). Quantitative reverse transcription polymerase chain reaction (RT-qPCR) was used to determine the expression levels of IL-6, IL-10, TNF-α, and IL-2sRα genes. According to the manufacturer’s instructions, total RNA was extracted from the LC of two individual samples using a Quick-RNATM Miccroprep kit (Zymo Research, CA, USA). Complementary DNA (cDNA) was synthesized using 2 μg of total RNA using iScriptTM cDNA Synthesis Kit (Bio-Rad, Hercules, CA, USA). Quantification was performed using Power SYBRTM Freen PCR Master Mix (Applied Biosystems, Whaltham, MA, USA) with the following protocol: two minutes at 95°C and for 40Lcycles of 15 s at 95°C and 1 min at 60°C using the 7500 Thermocycler (Applied Biosystems). Reactions were performed in two replicates on a 96-well plate. Raw Ct values obtained were used to normalize and calculate fold changes. The beta-actin gene was used as an endogenous control, and the primers employed are listed in Table 2.

**Table 2:**
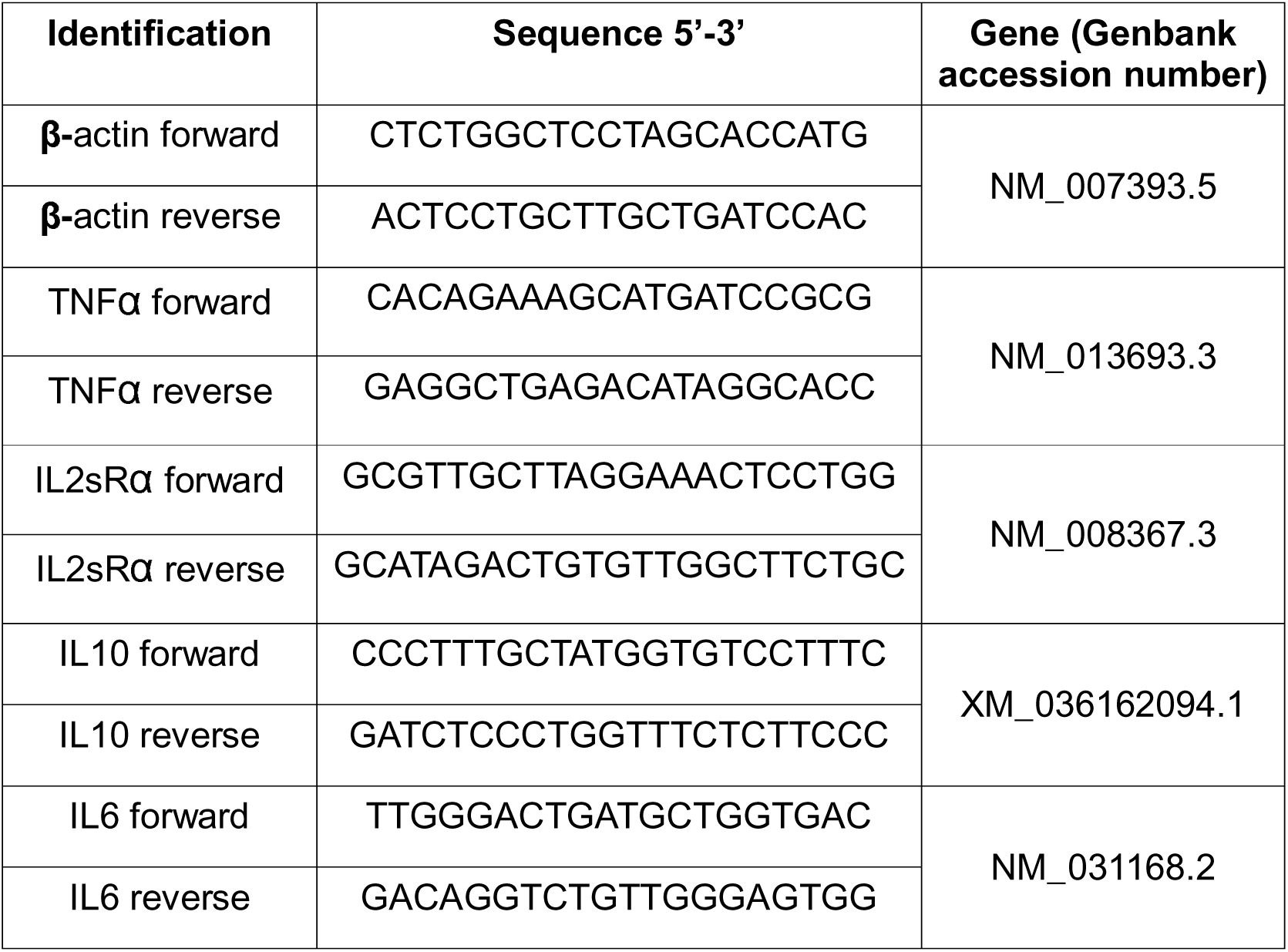
Primers used for RT-qPCR.

#### 2.1.6 Experimental protocols

##### Experiment 1: LC microglial activity to hypercapnic stimulus

Two groups of animals were used to investigate whether LC microglia is altered due to hypercapnia exposure. On the test day, all animals were first exposed to room air for 60 minutes to habituate them to the gas chamber and experimental conditions. Immediately after, one group remained with atmospheric air for 15 minutes (control), while the other inhaled 20% CO_2_ for the same period. The animals were then returned to their cages in the animal facility and remained there until euthanasia. Three different time intervals between the respiratory challenge and brain collection for morphological analyses were followed: 1 (n = 5 females and n = 4 males), 6 (n = 6 females and n = 6 males), or 24 hours (n = 5 females and n = 5 males) (Figure 1A).

##### Experiment 2: Systemic minocycline or clonazepam treatment on the behavioral and respiratory response to CO_2_

The effects of minocycline on the behavioral and respiratory responses to 20% CO_2_ were compared to those caused by the standard panicolytic drug clonazepam. Two experiments were performed. In the first, animals were injected with minocycline (40 mg/kg, n = 15) or vehicle solution (n = 17), and in the second with clonazepam (0.05 mg/kg, n = 9) or vehicle (n = 10). In both cases, the drugs were administered intraperitoneally (ip) for 14 days, at the same time of day (8 AM) throughout the treatment. One day after the last injection, animals were tested in the experimental setup (Figure 1B).

All animals were first exposed to normocapnia for 1 hour to habituate to the chamber and experimental conditions. Immediately after, with the animals still under room air conditions, respiratory parameters (see above in section 2.1.3) were recorded for the next 20 min (baseline scores). In the final 15 minutes, all animals were exposed to CO_2_ for behavioral analysis and a new respiratory assessment.

##### Experiment 3: Effect of systemic minocycline or clonazepam treatment on the plasmatic and LC cytokine levels

To assess the impact of systemic minocycline or clonazepam treatment on cytokines levels in the LC and plasma, the animals were divided into three groups, according to the drug treatment (i.p injections, 14 days): minocycline (40 mg/kg, n = 12), clonazepam (0.05 mg/kg, n = 6) or vehicle solution (n = 11) (Figure 1C). One day after the last injection, all animals were exposed to normocapnia for 1 hour. Following this habituation period, whereas one group of animals was exposed to room air for 15 minutes, the other was exposed to severe hypercapnia (20% CO_₂_) for the same period. Six hours later, the animals were decapitated, and blood was collected and immediately transferred into heparinized tubes. In addition, the brain was removed for LC extraction for RT-PCR analysis as described above.

#### 2.1.7. Statistical analysis

Statistical analyses were performed using the Statistical Package for the Social Sciences (SPSS) version 26.0. The normality of the distribution of the variables was tested using the Kolmogorov–Smirnov test. Almost all experiments included both male and female subjects. At least eight animals per group were included to achieve statistically significant results with an error rate of less than 5%. Sex was incorporated as a factor in analyzing microglial activity, behavior, respiratory function, and metabolic parameters. As no significant sex differences were detected in any of the evaluated parameters, data from male and female subjects were pooled for analysis. To reduce animal use in accordance with ethical guidelines and the principle of reduction, specific procedures (clonazepam administration and cytokine quantification) were performed exclusively on males. In experiment 1, the effects of hypercapnia exposure on LC microglial morphology at different time intervals were analyzed by ANOVA. In experiment 2, whereas the impact of minocycline x vehicle or clonazepam x vehicle on the behavioral responses to CO_2_ was analyzed by Student t-test, their effects on body temperature, respiratory, and metabolic were submitted to a two-way ANOVA, with drug treatment (minocycline x vehicle or clonazepam x vehicle) and gas condition (normocapnia and hypercapnia), as independent factors. In experiment 3, plasmatic and LC cytokine levels were analyzed by a two-way ANOVA with drug treatment (minocycline, clonazepam, or vehicle) and gas condition (normocapnia and hypercapnia) as independent variables. When appropriate, Tukey’s post hoc test was used for multiple comparisons. All data were presented as means ± S.E.M.

### 2.2 Human study

#### 2.2.1 Study design and participants

This was a randomized controlled trial that compared PD subjects (n = 49) taking minocycline or clonazepam. It was a single-center, outcome-assessor-blinded, parallel-group study at the Psychiatry Institute of the Federal University of Rio de Janeiro from January 2022 to June 2023. The intervention groups (40 women and nine men) received a pill of minocycline (100 mg/day; n = 23) or a pill of clonazepam (0,5 mg/day; n = 26) for 7 days before the CO2 challenge. The 7-day administration period was chosen to allow both medications to reach steady-state plasma concentrations, thereby ensuring consistent pharmacological effects during the CO_₂_ challenge, and due to the fact that prior research has demonstrated that a 7-day course of minocycline effectively achieves neuroprotective and anti-inflammatory effects (42,43).

Individuals were included in the PD group if they were (1) diagnosed with PD and (2) self-reported no lifetime psychiatric medication use in the last 7 days before CO_2_ experiments. Potential subjects were also excluded if they had uncontrolled cardiovascular, endocrinological, hematological, hepatic, renal, or neurological disease, autoimmune conditions, chronic infection (i.e., HIV, hepatitis B or C), a history of liver abnormalities, or evidence of infection within one month of screening, and their respective treatments, such as steroids, antiretroviral therapy, anti-inflammatory, or chemotherapy. These illnesses and therapies may represent a bias in the study’s interpretation. Figure 3 shows the schematic view of the experimental design.

**Figure 3:**
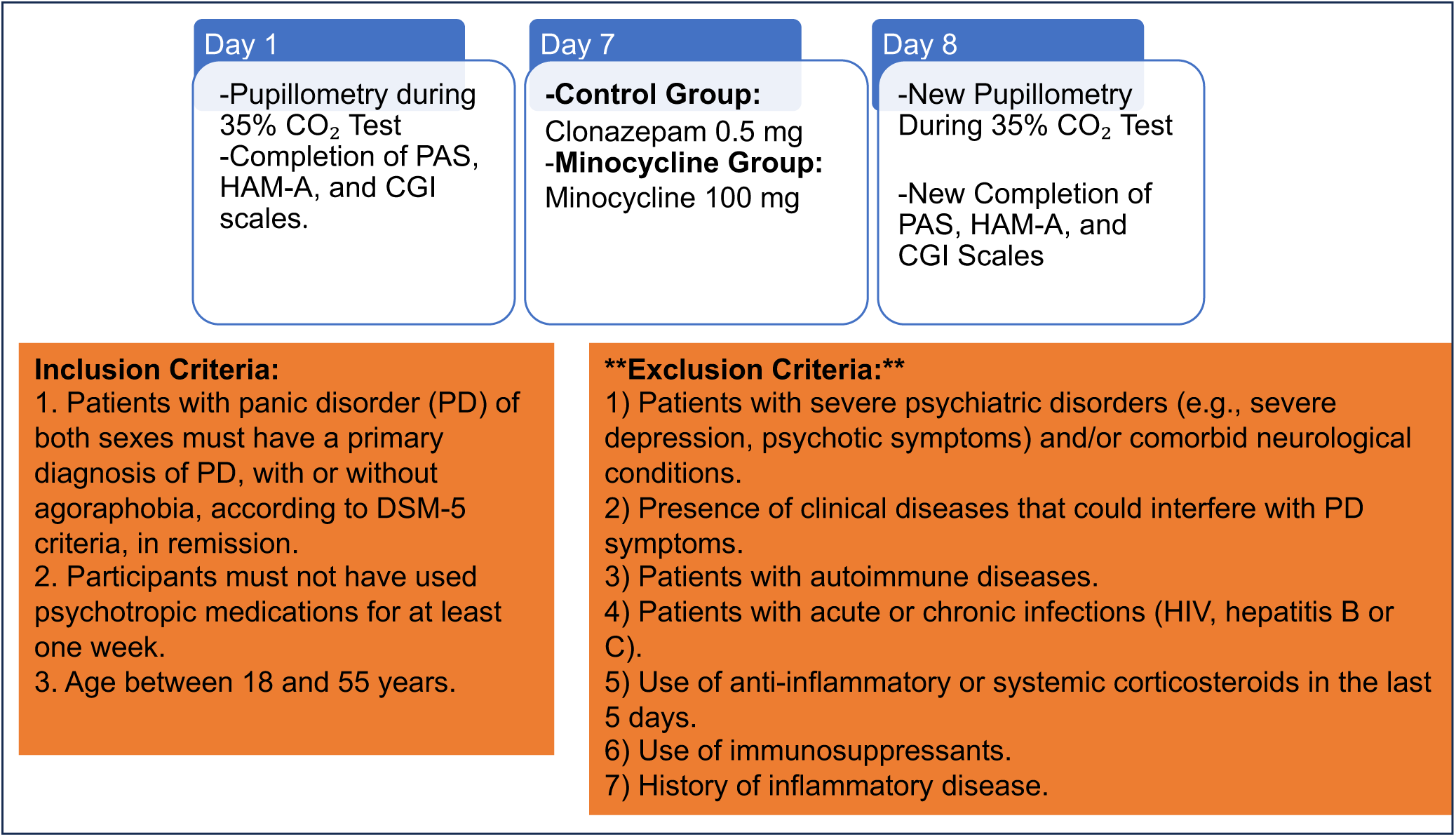
Schematic illustration of the experimental design for the human study.

#### 2.2.2 Clinical assessments and experiment

PD diagnosis was determined by a coordinated clinical interview based on the Diagnostic and Statistical Manual of Mental Disorders, Fifth Edition (DSM-5), administered by a trained psychiatrist or psychologist and independently confirmed by a research psychiatrist. Subjects inhaled 35% CO_2_ using a mask on two occasions: at the beginning of the study and after seven days of therapy with minocycline 100 mg/day or clonazepam 0.5 mg/day. To practice the CO_2_ challenge procedure and decrease any anticipatory anxiety, participants underwent a compressed air test inhalation procedure. After 30 minutes, they repeated the procedure using 35% CO_2_ gas inhalation. Patients were blinded to the two gas compositions: first, compressed air, and second, 35% CO_2_. Although the exact volume of air inhaled was not individually measured, all participants inhaled a standardized 35% CO_₂_ mixture with a fixed duration of inhalation, following established protocols demonstrating reproducible panic responses (44). This approach ensures consistency across participants and studies, facilitating comparison and reproducibility. A challenge was deemed successful when participants displayed characteristic physiological and subjective panic symptoms as per standardized criteria outlined in previous literature (44). The exposure occurred in a room with constant lighting in front of the computer, which was equipped with an eye tracker system to measure pupil diameter during the protocols. The primary outcome measures included physiological responses, subjective symptom ratings using validated scales, and cytokine levels. The scales were used to assess the severity and subjective experience of panic, complementing the objective physiological data.

The Hamilton Anxiety Rating Scale (HAM-A) and Panic and Agoraphobia Scale (PAS) were administered to all patients at baseline and after 7 days of treatment with the drugs to obtain measures of general psychopathology, and the Clinical Global Impression (CGI) scale was applied to evaluate total symptom severity. All assessments were made by psychiatrists trained in administering and scoring the assessments.

The height and weight of all participants were recorded to calculate their body mass index (BMI). Venous blood collections for interleukin measurement were carried out at baseline in all patients and after 7 days of therapy with minocycline or clonazepam, 30 minutes after the second CO_2_ test. The procedures were explained, and written informed consent was obtained from participants before inclusion in the study, which was approved by the research ethics committee of the Federal University of Rio de Janeiro (CAAE 55210222.2.0000.5263). This study was performed in accordance with the ethical standards of the Declaration of Helsinki.

A researcher without clinical involvement in the trial created the randomization sequence using Statistical Analysis Software (SAS) 9.3 (New Cary, North Carolina, USA). The randomization sequence employed a 1:1 allocation, with random permuted block sizes of 4 and 6. Upon the patient’s signing of informed consent and assent, authorized study personnel used a password-protected web-based system that implemented the randomization sequence. The system automatically assigns eligible patients to treatment groups upon entering the patients’ initials and details of stratification factors. Patients and clinicians who conducted outcome assessments were kept blinded to the treatment allocation.

#### 2.2.3 Pupil diameter

The pupillary diameter assessment tests were performed similarly to previous studies (45) and were performed on the baseline and after the intervention with minocycline or clonazepam. The experiments were conducted in a room with a constant ambient luminance and light intensity that did not exceed that of a typical 20-cycle fluorescent lamp. Pupillary diameter was continuously recorded throughout the entire CO_₂_ challenge procedure, starting from baseline (compressed air) and extending until the end of the 35% CO_₂_ inhalation, ensuring that both the onset and peak of the physiological response were captured for all participants. To standardize procedures, all participants inhaled the gas mixture for a fixed and validated duration. The participants sat 65 cm from a 21.5-inch computer monitor with a resolution of 1920 × 1200 (Dell ST2220Mb; Dell Corporation, Texas, USA). A Tobii Pro Nano Eye Tracker (Tobii AB Inc., Danderyd, Sweden) was attached to the monitor to record the pupillary diameter during testing. Tobii has excellent accuracy and precision with a high tolerance to head movements, and it was not necessary to restrain the participants’ heads with a stabilizer. Since this procedure was identical across participants and conditions, potential interference was equally distributed and unlikely to bias between-group comparisons. Before formal testing, each participant received a complete and detailed explanation of the test procedure.

#### 2.2.4 Quantification of cytokines

Blood was obtained in the morning (8 a.m. ± 1 h) in EDTA tubes through a catheter after participants had rested for at least 30 minutes (baseline samples). Blood was also collected in the morning (8 a.m. ± 1 h) in EDTA tubes after the CO_2_ experiment, which occurred 7 days after intervention with minocycline or clonazepam.

Blood was immediately centrifuged (1000 xg for 10 min), and plasma was collected and stored at −80 °C until the batch assay. Plasma samples were collected and stored in an ultrafreezer. Subsequently, the Quantification of IL-6, IL-10, TNF-α, and IL-2sRα was performed using an enzyme-linked immunosorbent assay (ELISA), following the manufacturer’s instructions. The detection limits for each cytokine are specified in Table 3.

**Table 3.** Cytokines Evaluated with Their Respective Detection Limits.

| Cytokine | Detection limit | Manufacturer | Catalog |
| --- | --- | --- | --- |
| BD OptEIA™ Set humanIL-6 | 4.7 pg/mL | BD | #555220 |
| BD OptEIA™ Set human IL-10 | 7.8 pg/mL | BD | #555157 |
| BD OptEIA™ Set human TNF-α | 7.8 pg/mL | BD | #555212 |
| BD OptEIA™ Set human IL-2sRα | 7.8 pg/mL | BD | #559104 |

#### 2.2.5 Statistical analysis

Statistical analyses were performed using the Statistical Package for the Social Sciences (SPSS) version 26.0. The normality of the distribution of the variables was tested using the Kolmogorov–Smirnov test. Comparisons of demographic variables were made through X2, Fisher’s, or Mann-Whitney U tests. Independent samples t-tests were used for clinical and biological parametric variables, and Mann-Whitney U tests were used for clinical and biological nonparametric variables. To evaluate the reduction in the severity of panic attacks, repeated measures analysis of variance (ANOVA) was used. The comparison of intragroup cytokine concentration in baseline and after the experiment was performed using the Wilcoxon test for related samples. A comparison of cytokine concentration between groups was performed using ANOVA. Multivariate linear regression models were performed to evaluate the effect of different types of treatment (clonazepam or minocycline) on cytokine levels. Statistical significance was set at p < 0.05, except on the Wilcoxon test, where the statistical significance was set at p < 0.001. All data were presented as means ± S.E.M.

## 3. Results

### Mice studies

#### Experiment 1 – Severe hypercapnia promotes morphological changes in LC microglial cells

The microglial analysis data showed a significant difference between the exposure (hypercapnia or normocapnia) and the time intervals (exposure versus time interval: F(2,25) = 81.573, p < 0.001; Figure 4; Table S1). Exposure to 20% CO_2_ resulted in a reduction in the arborization area of LC microglial cells, demonstrating a significant difference compared to normocapnia, as well as within the 6-hour time interval (t(10)= 13.570, p < 0.001, Figure 4A). Moreover, there were no significant differences observed in the arborization area between normocapnia and hypercapnia conditions across the 1-hour and 24-hour time intervals (hypercapnia 1-hour time interval: 894.5 ± 39.8 versus normocapnia 846.1 ± 35.6, p = 0.374; hypercapnia 24-hour time interval: 913.3 ± 32.4 versus normocapnia 879.3 ± 39.8). Morphological changes in cell body area revealed an increase under hypercapnic conditions within the 6-hour time interval (t(10)= 5.339; p<0.001, Figure 4B). No significant differences were detected between normocapnia and hypercapnia conditions across the 1-hour and 24-hour time intervals (hypercapnia 1-hour time interval: 12.8 ± 1.4 versus normocapnia 13.2 ± 1.2; hypercapnia 24-hour time interval: 11.4 ± 1.1 versus normocapnia 12.6 ± 1.4). Exposure to hypercapnia resulted in a decrease in the cell density of microglia cells after 6 hours (t(10)= 6.681; p < 0.001; Figure 4C). An increase in the morphological index was observed under hypercapnic conditions within the 6-hour interval (t(10)= 22.910, p <0.001; Figure 4D) and regarding the nearest neighbor distance (NND), an increase is observed under hypercapnic conditions compared to normocapnia within a 1-hour time frame (t(7)= 3.661; p= 0.008; Figure 4E). Figure 4F shows a representative photomicrograph of immunohistochemistry for Iba-1 in the LC region under normocapnic and hypercapnic conditions at different time intervals.

**Figure 4:**
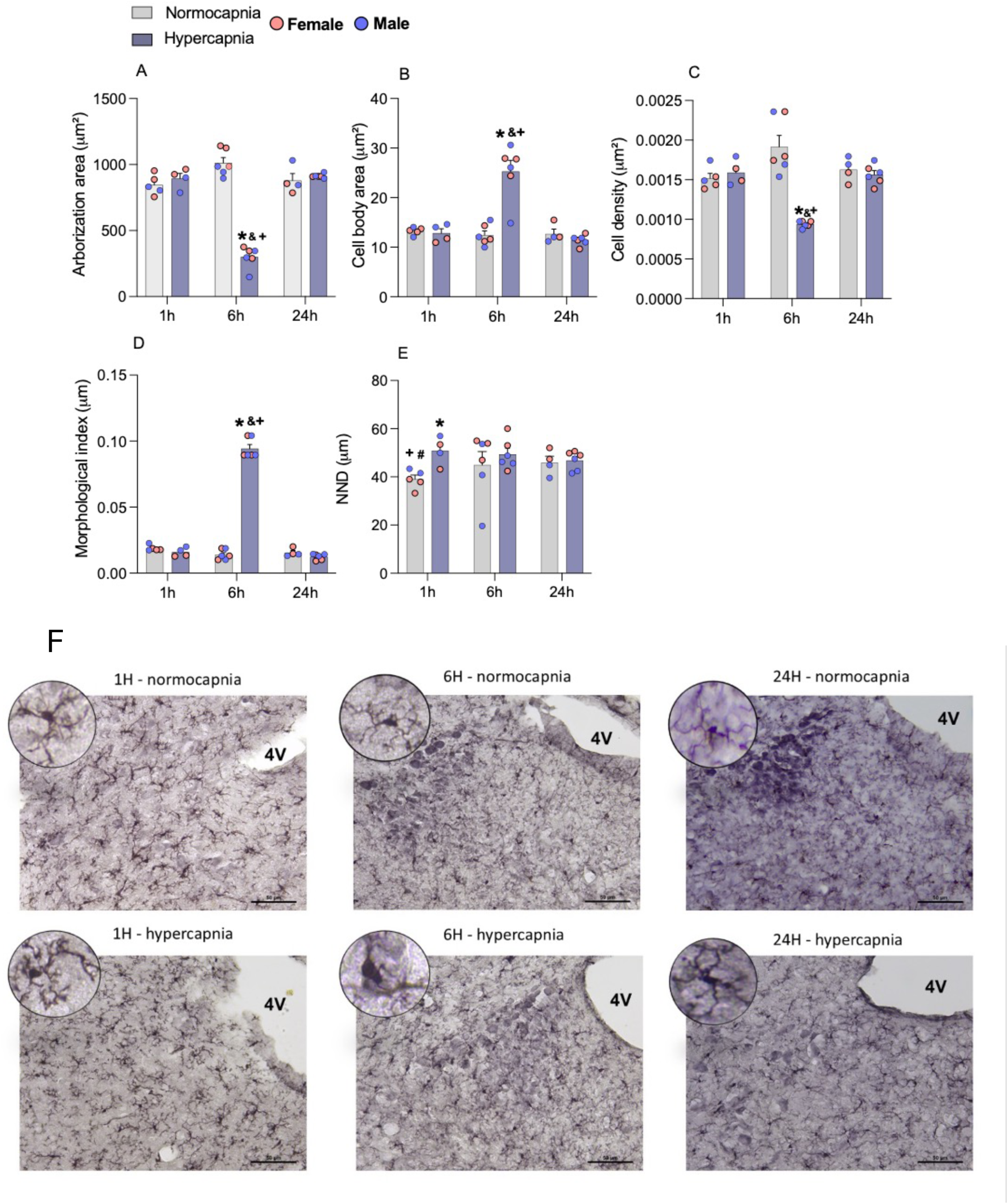
Effect of hypercapnic conditions on. **A**) arborization area, **B**) cell body area, **C**) cell density, **D**) morphological index, and **E**) nearest neighbor distance (NND) of microglial cells in the LC region analyzed over three time intervals (1 hour, 6 hours, and 24 hours) after the stimulus. **F**) Representative photomicrography of immunohistochemistry for Iba-1 in the LC region under normocapnic and hypercapnic conditions, at different time intervals. Scale is 50 µm. * Indicates a significant difference between the means of the normocapnia and hypercapnia groups; ^&^ indicates a significant difference between the 1 h interval; ^#^ indicates a significant difference between the 6 h interval; + indicates a significant difference between the 24 h interval.

#### Experiment 2 – Minocycline and clonazepam treatment decreased the expression of panic-related escape responses

Figure 5 illustrates the effects of minocycline and clonazepam on the behavioral responses exhibited by mice during CO_2_ exposure, as none of the defensive responses measured were detected while the animals breathed atmospheric air (Table S2). Figure 5A shows that chronic treatment with minocycline significantly decreases the number of jumpings [t(30)= 2.115, p < 0.05], without affecting the number of running episodes (Figure 5B) or the percentage of time in freezing (Figure 5C).

**Figure 5:**
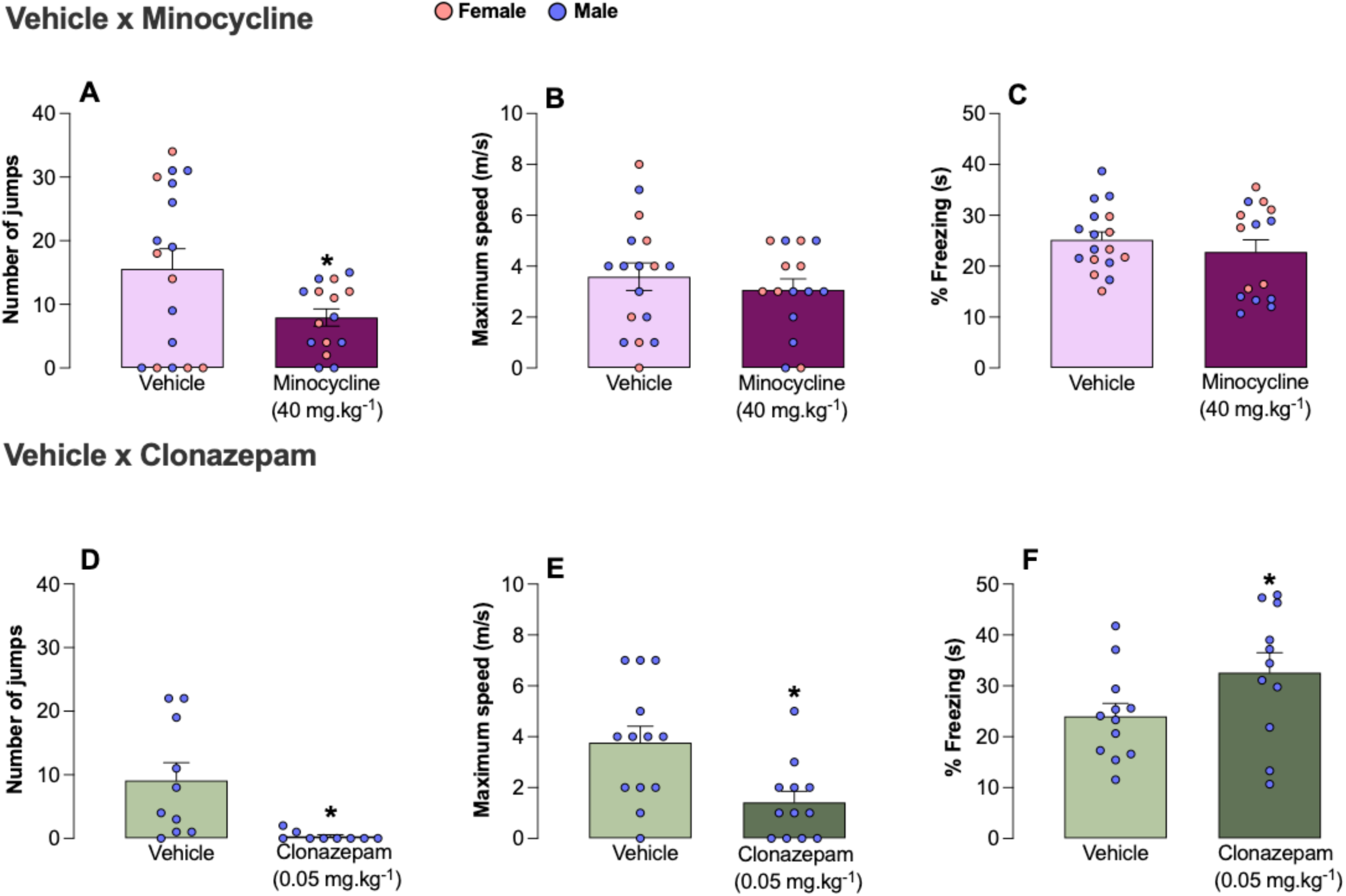
Effect of systemic treatment with vehicle, minocycline (40 mg/kg/day) or clonazepam on freezing. (**A, D**), number of jumps (**B, E**), and runs (**C, F**) in mice exposed to 20% CO_2_ challenge. *Indicates a significant difference between the means of the vehicle and treated group.

Treatment with clonazepam decreases both the number of jumpings (Figure 5D; t(17)= 2.938, p < 0.001) and runnings (Figure 5E; t(17)= 3.091, p < 0.05) and increases the percentage of freezing (Figure 5F; t(17)= 2.937, p < 0.05).

None of the treatments significantly altered locomotion in the experimental chamber (Supplementary Material – Figure S1).

As shown in Figure 6, during normocapnia, treatment with minocycline or clonazepam did not induce changes in the ventilatory response or tidal volume. However, minocycline did attenuate the respiratory frequency (f_R_ minocycline: 260.5 ± 9.1 versus vehicle: 299.0 ± 9.4, F(1,58)= 8.498, p < 0.001; Tables S6 and S7).

**Figure 6:**
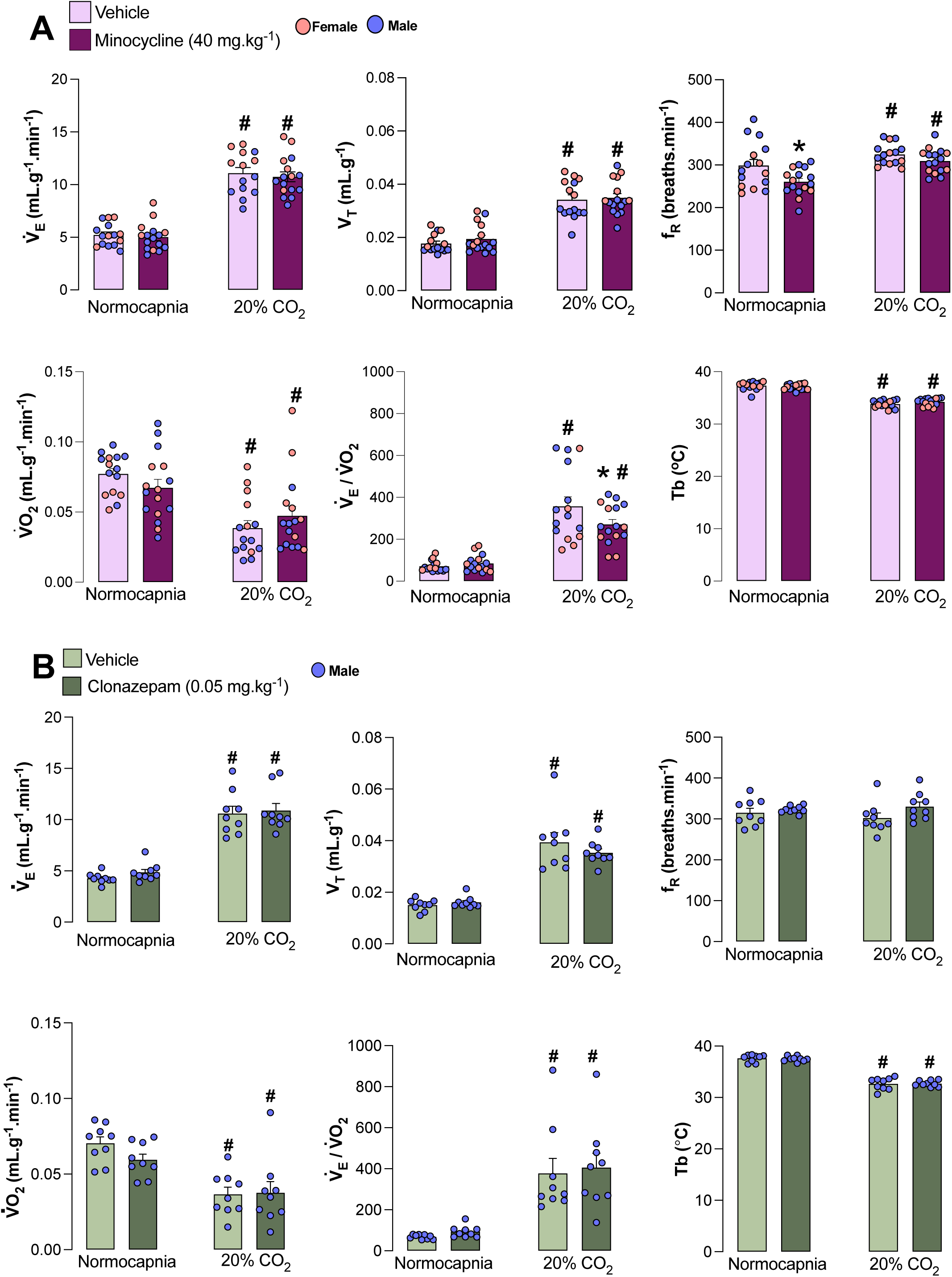
Effect of systemic treatment with minocycline (40 mg/kg/day) (**A**) or clonazepam (0.05 mg/kg/day) (**B**) on ventilation (V_E_), tidal volume (V_T_), respiratory rate (f_R_), oxygen consumption (VO_2_), respiratory equivalent (V_E_/VO_2_) and body temperature (Tb) under normocapnic and hypercapnic conditions (20% CO_2_). *Indicates a significant difference between the means of the vehicle and treated groups under the same condition. #Indicates a significant difference between the means of the same group between normocapnic and hypercapnic conditions.

Regarding exposure to the hypercapnic challenge, 20% CO_2_ increased the ventilatory response compared to the normocapnic group (F(1,58)= 206,023, p < 0.0001), in all groups. This increase in ventilatory response during hypercapnia was driven by an increase in V_T_ in all groups (F(1,58)= 122,995, p < 0.001) and respiratory frequency for minocycline and its vehicle (Figure 6; F(1,58)= 16.126, p < 0.001). No significant differences were found among groups in the ventilatory parameters under CO_2_ exposure (Tables S3 and S4).

During normocapnia, no difference in VO_2_, V_E_/VO_2,_ or Tb was observed among groups (Figure 6; Tables S5 and S6). CO_2_ exposure caused a reduction in VO_2_ in all groups (F(1,58)= 26.770, p < 0.001) and an increase in V_E_/VO_2_ relation (F(1,58)= 83.736, p < 0.001). Exposure to high concentrations of CO_2_ resulted in a significant reduction of body temperature (Tb) (F(1,58) = 17.636, p < 0.001) in all groups, and this effect was not influenced by the treatment (Tables S3 and S4).

#### Chronic minocycline or clonazepam did not change cytokine levels during normocapnia and hypercapnia

The data regarding the baseline normocapnic and CO_2_ conditions of animals treated with vehicle, minocycline, and clonazepam are shown in Figure 7. As observed, only plasma IL-10 levels were affected by chronic treatment with minocycline and clonazepam (F(2,61)= 3.717, p<0.05) (Figure 7). Plasma IL-6 is also affected by exposure (F(1,62)= 13.450, p<0.05), with no differences among treatments (Figure 7). The other cytokines, both in plasma and in the LC, were unaffected by the treatment under any of the tested conditions (Figure 7 and Tables S5 and S6).

**Figure 7:**
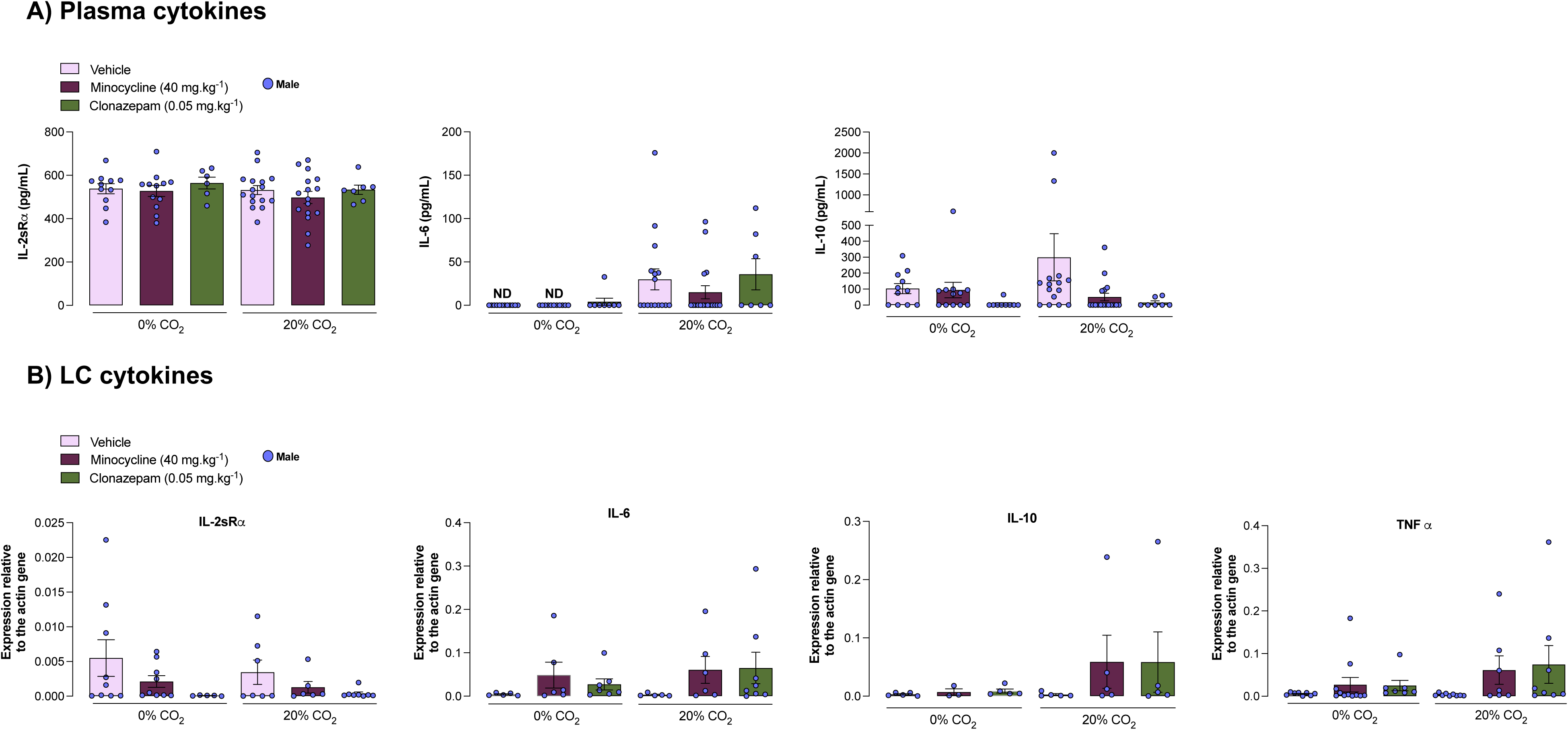
Effect of systemic treatment with minocycline (40 mg/kg/day) or clonazepam (0.05 mg/kg/day) on plasma. (**A**) and *Locus coeruleus* cytokines (**B**) under normocapnic or hypercapnic conditions (20% CO_2_). + Indicates a significant difference between the means of the same group between normocapnic and hypercapnic conditions. * Indicates a significant difference between the means of the vehicle and treated groups under the same condition.

### Human study

#### Demographic and clinical differences between minocycline and clonazepam groups

The sample was composed of 49 individuals (40 women and 9 men) with a PD diagnosis. The sample was successfully matched, and there were no significant differences between groups regarding sex (X2 (1) = 1.723; p= 0.273), ethnicity (X2 (2) =0.428; p= 0.914), religion (X2 (4) =3.686; p= 0.512), marital status (X2 (3) = 3.238; p= 0.336), salary (U=289.0; p=0.838) and education (U= 295.5; p=0.943). Only age showed a statistically significant difference (U= 159.0; p=0.005). The results are shown in Table 4. Regarding the pre-treatment interview about symptoms of agoraphobia and the risk of suicide, no difference was found among the evaluated patients (Figure 8).

**Figure 8:**
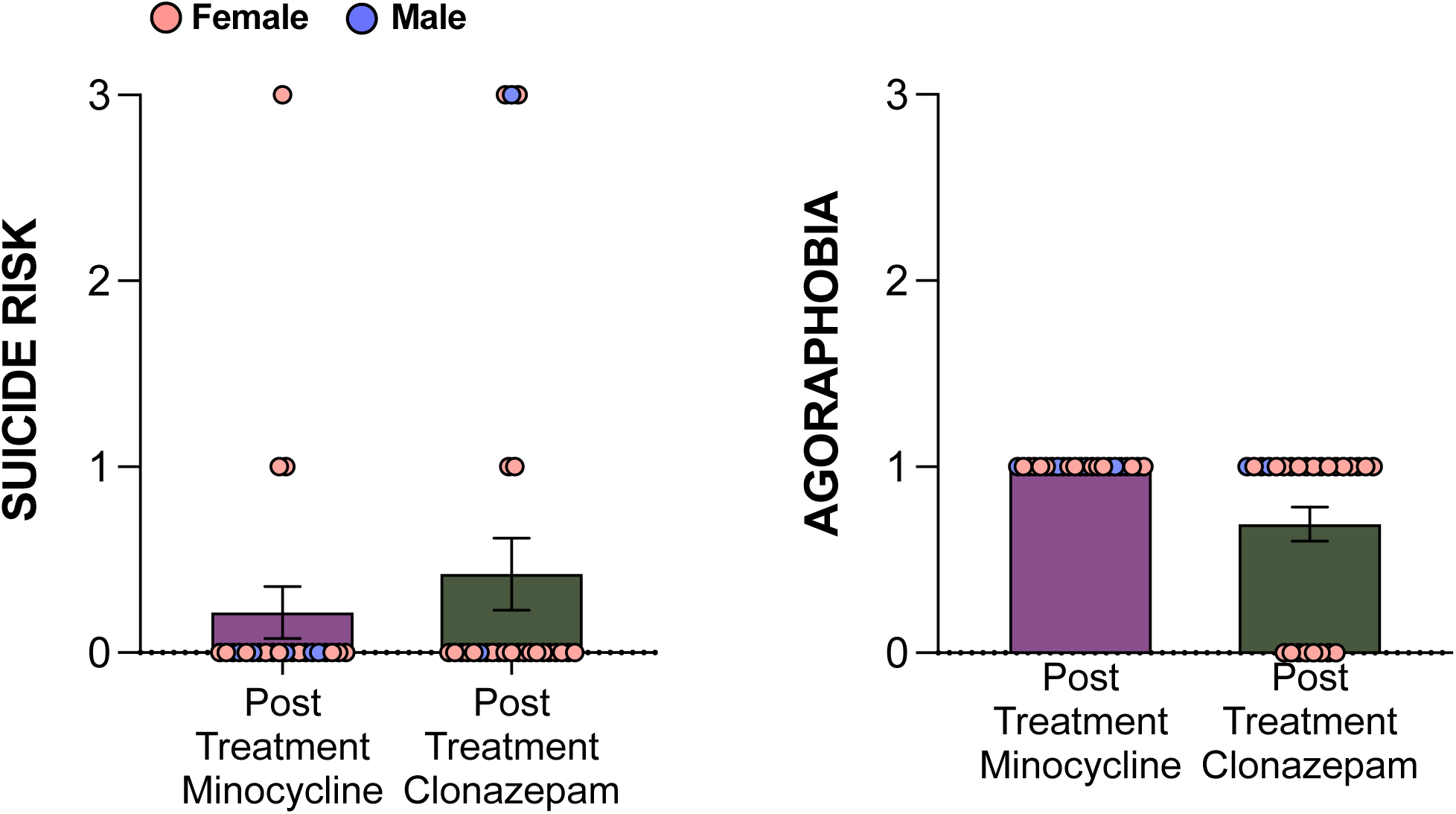
Effect of minocycline or clonazepam treatment on panic disorder patients in the parameters of agoraphobia and suicide risk.

**Table 4:**
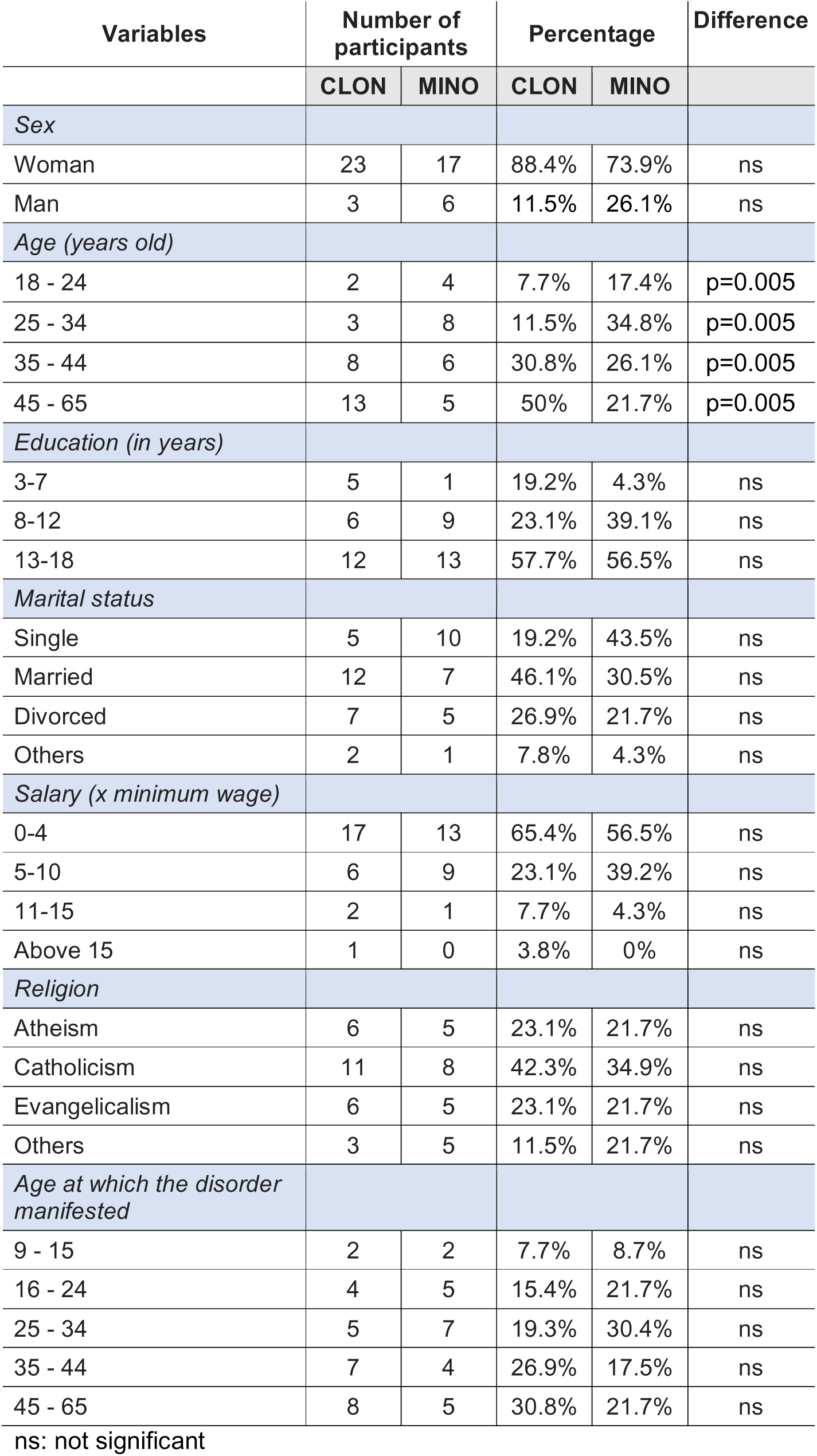
Demographic and clinical differences 1166 between minocycline group and clonazepam controls.

The times PAS1 and PAS3 in the graphs indicate 7 days before treatment with clonazepam or minocycline (PAS1) and 7 days after treatment with clonazepam or minocycline (PAS3). The analysis of clinical data measured by PAS, HAMA and CGI demonstrated that PAS (U= 112.5; p=0.000), (U= 132.0; p=0.001) and HAMA scales (U= 285.0; p=0.777), (U= 254.5; p= 0.369) were constant pre and post-therapeutic approaches, and were statistically significant. However, the CGI scale showed a statistical difference between the groups only in the post-CGI test [(U= 275.0; p=0.589), (U= 187.5; p=0.007)] (Figure 9). The measurements obtained from the eye tracker were statistically significant between the groups at both moments of data collection: eye track right eye baseline (U= 194.0; p= 0.004) and post intervention (U= 205, 0; p= 0.021); and eye track left eye baseline (U= 177.5; p= 0.006), and post-intervention (U= 205.0; p= 0.057) (Figure 10).

**Figure 9:**
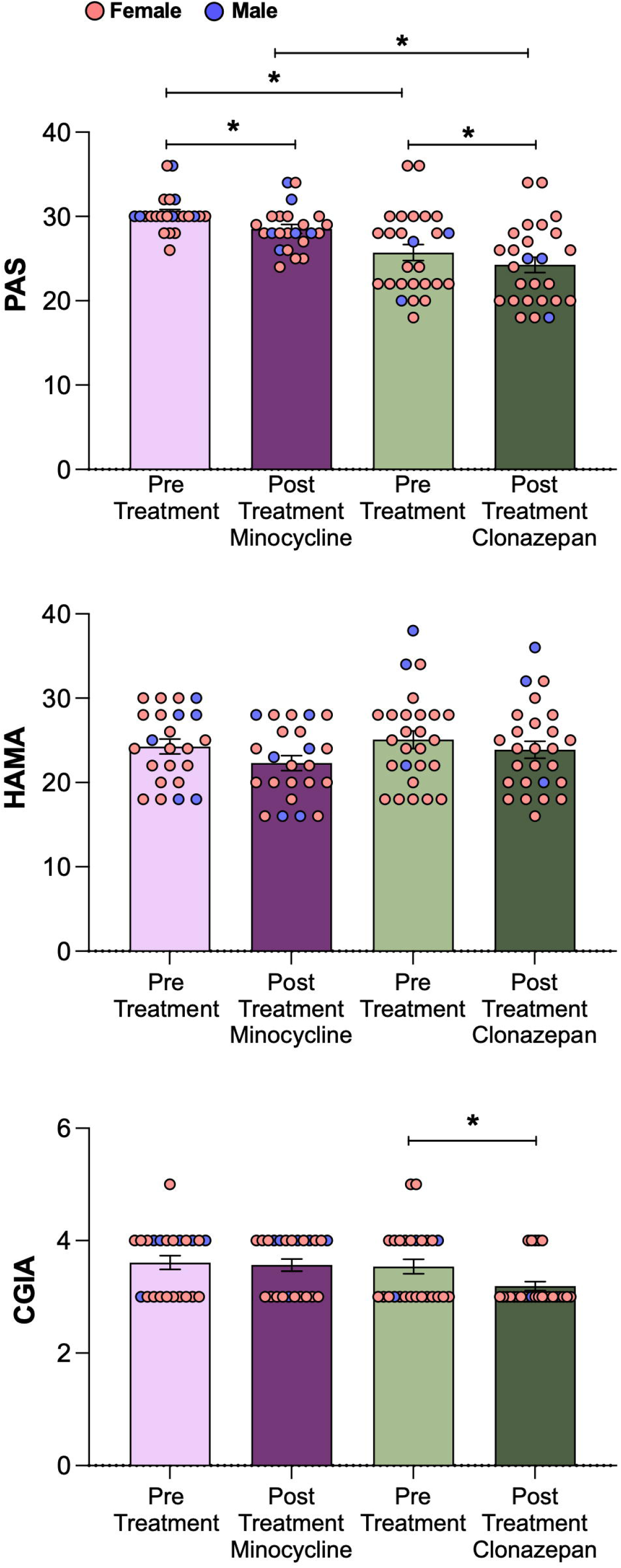
Effect of minocycline or clonazepam treatment on panic disorder patients on the Hamilton Anxiety Rating (HAM-A), Panic and Agoraphobia Scale (PAS), and Clinical Global Impression (CGI) scales. *Indicates a significant difference among the SEMs of the groups.

**Figure 10:**
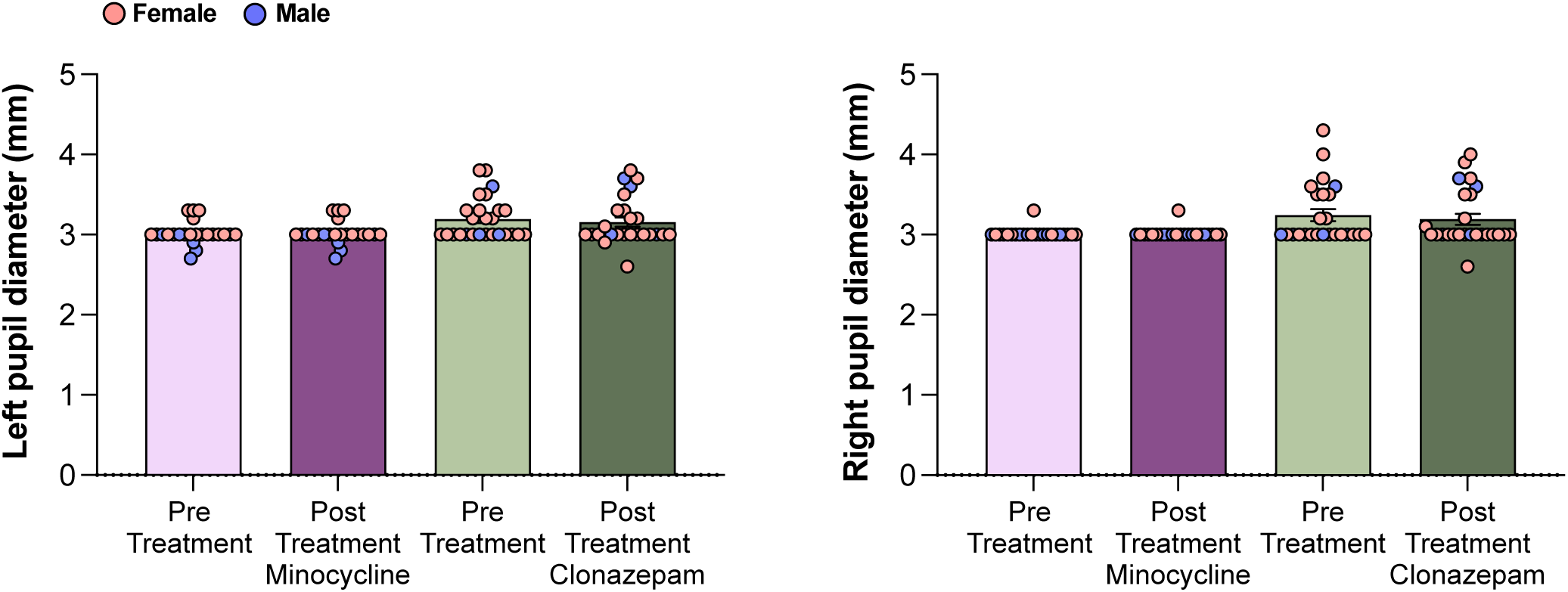
Effect of minocycline or clonazepam treatment on pupil diameter in panic disorder patients.

To evaluate the reduction in the severity of panic attacks, repeated measures analysis of variance (ANOVA) was used. The results demonstrated that minocycline and clonazepam reduced attacks over time F (1, 47= 94.268; p= 0.000)]. There was no statistical difference between the groups regarding drug therapy F (1, 47= 1.159; p= 0.287). Finally, to rule out the influence of sex on the results found in PAS in the baseline and after drug treatment, an analysis of covariance (ANCOVA) controlled by sex was carried out F (2, 46= 8.632; p= 0.721), F (2, 46= 7.263; p= 0.995).

#### Intra and intergroup cytokine analysis and panic severity symptoms

The comparison of intragroup cytokine concentration in the baseline and after 7 days of treatment was performed using the Wilcoxon test for related samples. The minocycline group demonstrated a statistically significant result regarding IL-2sRα (pre-treatment vs post treatment: F(3,94)= 35.190, p < 0.0001); IL-6 (pre-treatment vs post-treatment: F(3,94)= 8.020, p < 0.0006), IL-10 (pre-treatment vs post treatment: F(3,94)= 5.762, p < 0.05) and TNFα (pre-treatment vs post treatment: F(3,94)= 7.220, p < 0.001). The clonazepam group demonstrated a statistically significant result regarding only IL-2sRα (pre-treatment vs post-treatment: F(3,94)= 35.190, p < 0.0001). The cytokine concentration between groups was compared using analysis of variance (p < 0.005), and the results are presented in Figure 11.

**Figure 11:**
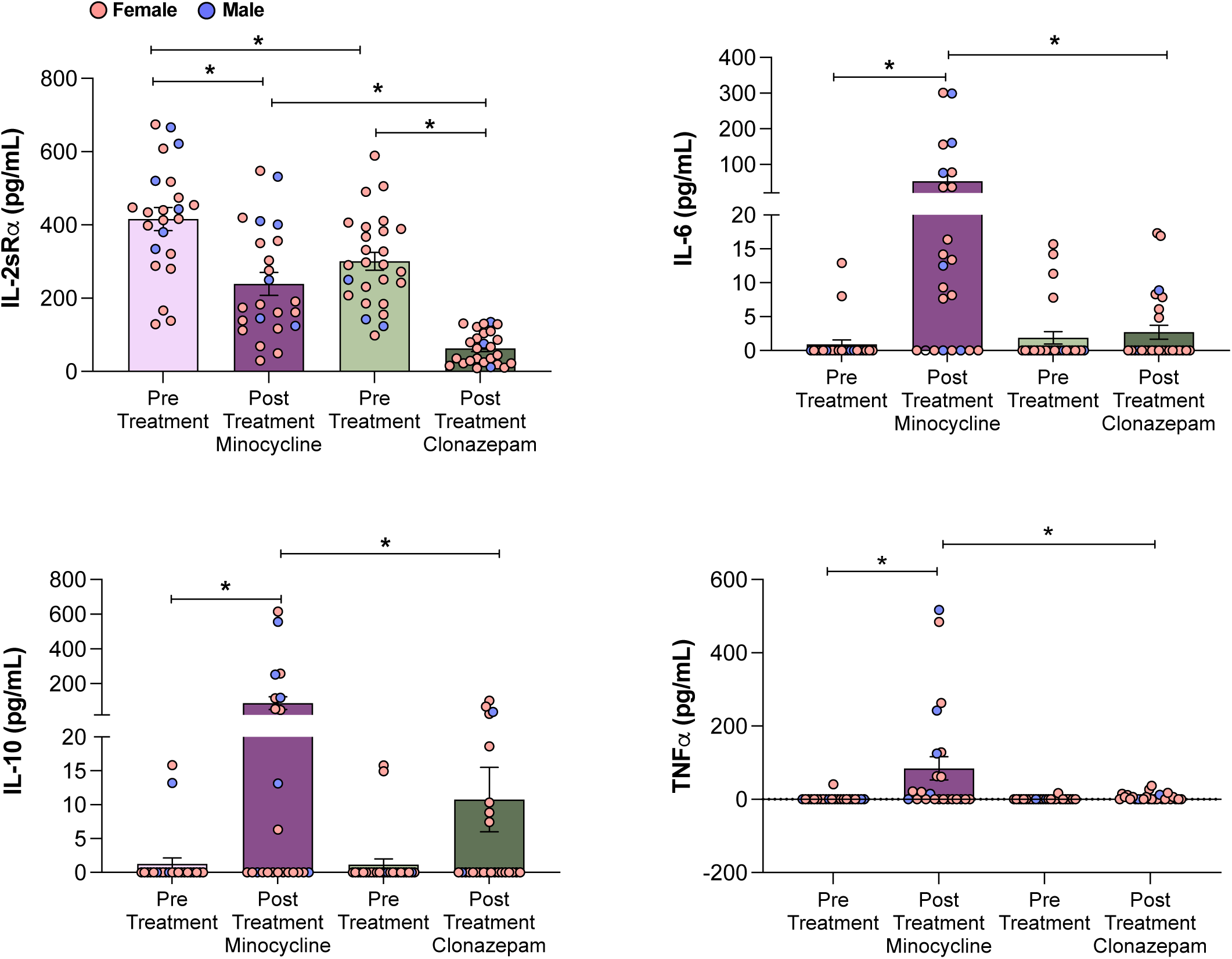
Evaluation of cytokine production in panic disorder patients after treatment with minocycline or clonazepam. *Indicates a significant difference among the means of the groups.

## 4. Discussion

Increasing evidence has suggested that microglial cells play a key role in the neural responses to internal threats, such as elevated levels of CO_2_, which are relevant to the pathophysiology of panic (22,46,47). Here, we show that acute hypercapnia leads to microglial activation in the LC, a region implicated in anxiety and panic-related behaviors. In addition, using a relevant animal model and a clinical sample of patients with PD, we compared the effectiveness of two distinct types of medications in treating panic attacks: minocycline, an antibiotic with anti-inflammatory properties, and clonazepam, a commonly used comparator in experimental and clinical studies of PD. In both models, treatment with minocycline or clonazepam was associated with reductions in panic-related behaviors and symptoms, suggesting that minocycline may have supportive therapeutic potential in panic disorder.

Although the morphological microglial approach may be considered outdated, it offers valuable insights into microglial characterization. We acknowledge that while IBA-1 expression and cytokine profiling are practical methods for assessing microglial activation, it is important to recognize that microglial morphology and classification are dynamic and continually evolving fields, as recently reviewed (48). Microglia have traditionally been categorized into M1/M2 phenotypes, but they exist on a spectrum with various transitional and mixed states influenced by local environmental factors. Recent approaches favor multidimensional and transcriptomic analyses, which may affect how activation and phenotype states are interpreted.

Our results showed microglial activation 6-, but not 1– or 24 hours, after a 15-minute exposure to 20% CO_2_. By analyzing morphological changes, such as the arborization area and cell body size, we identified that microglia were in their activated state. This conclusion is supported by the unbranched morphology and increased cell body size observed during microglial activation (49). Cell density, calculated by the total number of cells in the total area analyzed, also decreased in mice exposed to hypercapnia. Interestingly, a previous study found no evidence of microglia activation or development of a phagocytic phenotype until 6 hours after brain tissue sectioning (50). Supporting our findings, Rowe et al. (51) reported that microglia activation and pro-inflammatory cytokine expression occur only 5–6 hours after brain injury. Similarly, microglia in hippocampal slices did not exhibit morphological signs of activation until 12 hours after sectioning (49). Consistent with this observation, the transition from ramified to amoeboid microglia is not immediate but requires the near-complete retraction of existing branches before the cells display protrusive motility and locomotion (49). In alignment with these findings, our data demonstrate that exposure to high concentrations of CO_2_ activates microglia only after a 6-hour delay following stimulation.

While microglial activation was observed only 6 hours after CO_₂_ exposure, chronic minocycline treatment may have primed microglia earlier, subtly influencing circuits involved in panic responses. Rapid neuronal, chemosensory, and autonomic pathways mainly mediate the effects of CO_₂_ on ventilation and behavior. In contrast, microglial activation likely represents a delayed neuroimmune response to intense neuronal activity and fluctuations in pH and CO_₂_, contributing to synaptic remodeling or cytokine release rather than initiating panic behaviors. However, early microglial modulation by minocycline could have affected stress responses and shaped behavioral outcomes. Exploring these temporal dynamics with time-resolved markers and functional assays is an important direction for future research.

Our results also showed that marked ventilatory, metabolic, and behavioral changes in mice were observed during CO_2_. Accordingly, inhalation of this gas by mice elicits different behavioral responses ranging from freezing and avoidance at lower concentrations (22,52–55) to escape-like reactions (e.g jumps and running episodes) at higher ones (15,39,55). In our study, inhalation of 20% CO_2_ caused an increase in jumps and running episodes in both males and females, suggesting that the animals are facing a highly aversive situation, resembling the onset of a panic attack.

Experiment 2 also showed that minocycline treatment (40 mg/kg for 14 days) reduced the expression of panic-related behaviors to CO_2_, giving support to the potential role of this drug as a panicolytic agent. In the current study, we observed that minocycline reduced by approximately 41% the number of jumps, while clonazepam (0.05 mg/kg, 14 days) totally blunted this response and decreased the running episodes. Benzodiazepines are widely used in the treatment of PD and have been reported to reduce reactivity to CO_2_ inhalation in patients with this condition (56,57). The anti-escape effects observed with clonazepam and minocycline occurred at doses that did not nonspecifically affect locomotion, as assessed prior to CO_2_ exposure. Therefore, these findings indicate that clonazepam and minocycline did not cause significant locomotor impairment, supporting the idea that their effect on reducing escape responses is due to a specific anxiolytic action on panic-like behaviors rather than sedation or motor suppression. Minocycline has been shown to alleviate stress-induced anxiety in rodent models significantly (58–60). Studies suggest that this effect is mediated by the drug’s capacity to inhibit microglial activation, attenuate neuroinflammation, and modulate the release of pro-inflammatory cytokines, factors commonly associated with stress-related neural and behavioral alterations. However, in our study with mice, we did not observe significant differences in cytokine levels in either the plasma or the locus coeruleus. The high variability in the data, likely due to individual differences in inflammatory responses, may have masked potential treatment effects.

Although minocycline is well known for its anti-inflammatory activity, particularly via the suppression of microglial activation, its anxiolytic effect might not be explained entirely by such mechanisms. Indeed, modulation of microglia and anti-anxiety may be coexistent yet independent mechanisms. There are direct neurophysiological effects with minocycline as well. For instance, it is capable of modulating neuronal excitability by modulating ion channel activity in hippocampal neurons (61), dorsal root ganglion (DRG) neurons (62) and in substantia gelatinosa neurons (63). These mechanisms suggest that minocycline might decrease anxiety-related neural activity by direct effects on neurons independent of its immunomodulatory actions. Thus, the anxiolytic effects observed may be the result of a combination of its neuroprotective and neuromodulatory effects and not merely through inhibition of microglia. Therefore, further studies with larger sample sizes or more sensitive inflammatory markers may be necessary to clarify the relationship between cytokine modulation and behavioral outcomes.

Under normocapnic conditions, minocycline treatment did not alter the ventilation response or metabolic rate. However, the drug significantly attenuated the respiratory frequency, which was not observed with clonazepam, suggesting potential modulatory effects under baseline conditions, which may be considered beneficial a priori, as a baseline reduction in f_R_ could attenuate increases in this variable during respiratory challenge conditions. Similarly, a previous study using rhythmic slices containing the pre-Bötzinger complex demonstrated that minocycline treatment (30 μM) decreased f_R_ (64). According to the authors, microglia may be continuously releasing excitatory modulators (65), which may play a role in sustaining respiratory rhythm generation (66–68). Therefore, inhibiting or depleting microglia could attenuate this tonic excitatory modulation, leading to a reduction in the generation of respiratory rhythm (66–68).

In the current study, the decrease in respiratory frequency observed after minocycline treatment did not significantly affect minute ventilation under normocapnia or ventilation during high CO_₂_ levels, indicating that other compensatory mechanisms maintain overall ventilation at baseline. A trend toward a reduction in VO_2_ was observed in animals treated with minocycline under room air conditions, which could explain the lack of change in the respiratory exchange ratio (V_E_/VO_2_). This suggests that the change in respiratory frequency alone is unlikely to impact ventilation during a panic attack or heightened CO_₂_ exposure, as compensatory adjustments sustain adequate ventilation.

During hypercapnia, exposure to 20% CO_2_ led to an increased ventilatory response in all groups. Minocycline-treated animals attenuated the hypercapnic hyperventilatory response (V_E_/VO_2_), likely due to a combination of decreased V_E_ and increased VO_2_. Although these changes (V_E_ and VO_2_) were not statistically significant, the findings suggest a potential moderating effect of minocycline on hyperventilatory responses during hypercapnia. A previous study showed that systemic injection of minocycline in rats attenuated the ventilatory response to hypoxia (69). Another study also reported that microglial inhibition affects respiratory activity since intracisternal injection of minocycline reduces the ventilatory response to hypoxia (64). Furthermore, minocycline also decreased 24-h ventilatory acclimation to hypoxia due to blocking the continuous increase in tidal volume (70). However, the exact mechanisms involved in the modulation of brain respiratory areas by microglia are unclear but probably involve inflammatory mediators and neurotransmitters in the ventral respiratory column.

As shown by our clinical study, both minocycline and clonazepam treatments were associated with reductions in the severity of panic attack symptoms. However, our trial was not powered to establish non-inferiority or equivalence between treatments. Therefore, these findings provide preliminary, hypothesis-generating evidence suggesting that minocycline may have beneficial effects, which should be further investigated in larger randomized controlled trials. Additionally, our data demonstrated a significant improvement in anxiety severity with both drugs, as assessed by the PAS scale.

Selective serotonin reuptake inhibitors (SSRIs) and benzodiazepines are medications with proven efficacy in treating panic disorder. Interestingly, it has been shown that 50% of patients with PD continue to experience anxious symptoms despite pharmacotherapy (71), prompting the search for new therapeutic approaches. Additionally, benzodiazepines may be associated with specific risks such as dependence, tolerance, or sedation (72).

Patients with longstanding PD show alterations in circulating peripheral cytokine levels and hyperactivation of their immune system (73), making minocycline treatment an effective alternative to reduce the disorder’s symptoms as it is an anti-inflammatory medication. Minocycline is a highly lipophilic molecule that is rapidly absorbed orally and can cross the blood-brain barrier (74). It has been used in humans for over 30 years and is considered safe and well-tolerated at doses up to 200 mg/day, even with prolonged use. Despite its side effects (nausea, gastrointestinal irritation, dizziness, etc.), these usually regress after discontinuation, providing a reassuring safety profile (75). It has been frequently used as a neuroprotective and microglial inhibitory substance, limiting neuroinflammation and oxidative stress (76). Preclinical studies using minocycline to treat psychiatric disorders suggest that treatment with the drug attenuates anxiety and cognitive impairment in mice and rats (77). Our study showed no significant differences in reported side effects between the minocycline and clonazepam groups. Although our study did not evaluate combined treatment, it is conceivable that minocycline’s anti-inflammatory and neuroprotective actions could complement the anxiolytic effects of clonazepam. Future studies could explore whether concurrent administration may enhance efficacy or allow for lower benzodiazepine dosages, potentially reducing risks associated with long-term benzodiazepine use. However, such possibilities remain speculative at this stage and require dedicated investigation in preclinical and clinical settings

However, pupillometry testing did not indicate significant differences between treatments. Interestingly, the right and left pupil diameters differed between groups at both data collection times (pre– and post-treatment). Nevertheless, despite being considered different, they still fall within the normal range of 2 to 5 mm. Although pupillometry is a non-invasive and accessible technique, its main limitation lies in the lack of specificity regarding the spatial location of brain responses. Therefore, it is important to integrate pupillometry with real-time neuroimaging tests to improve spatial and temporal accuracy. Combining these tests would allow for investigating and identifying brain areas involved in the underlying mechanisms of panic attacks.

Previous study has demonstrated a significant increase in the inflammatory mediators such as IL-2R, accompanied by a decrease in the anti-inflammatory molecule IL-10, in nonmedicated PD patients (73). In the current study, significant differences in cytokine levels were observed when comparing pre-treatment and post-treatment groups with both minocycline and clonazepam. Pre-treated patients exhibited elevated levels of IL-2sRα and reduced levels of IL-10, consistent with findings from previous studies (46,47,73). Interestingly, a decrease in the expression levels of the pro-inflammatory interleukin receptor, IL-2sRα, and enhanced levels of the anti-inflammatory interleukin, IL-10, was noted in patients treated with minocycline and clonazepam. Even though the IL-6 and TNFα levels were higher when comparing pre-treatment and post-treatment with minocycline groups, our study did not identify a significant correlation for proinflammatory cytokines, such as IL-6 and TNF-α, in the clinical presentation of PD. It is important to note, however, that most prior findings are based on comparisons with healthy controls or patients experiencing acute exacerbations. Proinflammatory cytokines are more commonly associated with the severity of psychiatric disorders (47,73,78), whereas the absence or reduction of such associations has been less frequently explored. As a pleiotropic cytokine, the increase in IL-6 levels, which plays both proinflammatory and anti-inflammatory roles, requires further investigation. However, there may be a correlation with the increased levels of IL-10. Furthermore, IL-6 can exhibit different modes of expression, such as its release by muscle tissue, which imparts an anti-inflammatory effect. TNF-α, similarly to IL-6, is a proinflammatory cytokine; however, there have been divergent results regarding this cytokine in individuals with PD compared to healthy controls (46,47,73).

In conclusion, our research has shown that targeting microglia cells with minocycline can effectively reduce the panic-related response to CO_2_ in both humans and mice. These findings suggest a promising therapeutic approach for treating panic disorders. Moreover, the insights gained from this study on the pathophysiology of panic attacks could pave the way for the development of more precise and effective treatments for panic disorders.

## Data availability

The datasets generated and/or analyzed during the current study are available from the corresponding author on reasonable request.

## Acknowledgments and Disclosures

This work was supported by Fundação de Amparo à Pesquisa do Estado de São Paulo (FAPESP; 2020/01702-2 to LHG and 2021/04143-7 to BFGO) and Conselho Nacional de Desenvolvimento Científico e Tecnologico (CNPq; 302991/2022-0 to LHG).

BFGO, LAQ, AEN and LHG were responsible for the experimental idea and design; BFGO, LGAP, ATF, EMF, MER, BVBR, KCO, MMSB, CMM, TMSM, AIM and LHG were responsible for performing experiments, data analysis and figures of mice study; LAQ, FDG, NH and AEN were responsible for performing the human trial and data analysis and figures; BFGO, LAQ, ATF and LHG were responsible for manuscript writing. MMSB, CMM, TMSM, AIM was responsible for the interleukins (IL) analysis and figures and writing of the methods and results about IL. BFGO, ATF and MER performed the cell counts and ran analysis for the cell counts. LGAP helped with the respiratory experiments; BFGO and ATF performed the behavioral tests, and ATF analyzed the behavioral experiments and wrote the results section. KCB, HZJ, AIM, AEG and LHG mentored and edited the manuscript.

## Ethics declarations

### Ethics approval and consent to participate

All the experiments with mice were conducted with the approval of the local College of Agricultural and Veterinary Sciences Animal Care and Use Committee (CEUA-FCAV-UNESP-Jaboticabal; Protocol: n°11.794).

The procedures were explained, and written informed consent was obtained from participants prior to inclusion in the study, which was approved by the research ethics committee of the Federal University of Rio de Janeiro (CAAE 55210222.2.0000.5263). This study was performed in accordance with the ethical standards of the Declaration of Helsinki.

