## Supplementary material for "Minocycline attenuates panicogenic responses in a CO_₂_-induced panic attack model: a translational approach": Suplementary material

### SUPPLEMENTARY INFORMATION

#### SUPPLEMENTAL RESULTS

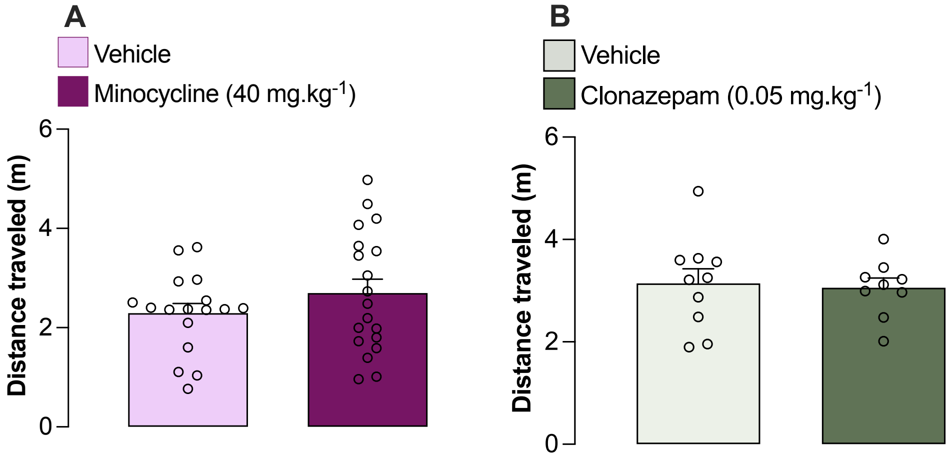


**Figure S1**: Locomotion activity of mice treated for 14 days of 40 mg/kg/day of minocycline (A) or 0.05 mg/kg/day of clonazepam (B) during room air conditions. Minocycline: $t(34)= 1.168$, $p= 0.251$; Clonazepam: $t(17)= 0.241$, $p= 0.812$

**Table S1:** Results of two-way ANOVA statistical analyzes for microglia analysis in the LC over three-time intervals in normocapnia or hypercapnia condition.

| Microglia analysis | | | | | |
|---|---|---|---|---|---|
| | **Arborization area** | **Cell body area** | **Cell density** | **Morphological index** | **NDD** |
| Exposure effect | $F(1,25)= 52.27$<br>$p < 0.001$ * | $F(1,25)= 12.91$<br>$p < 0.001$ * | $F(1,25)= 20.67$<br>$p < 0.001$ * | $F(1,25)= 241,101$<br>$p < 0.001$ * | $F(1,25)= 4.334$<br>$p <0.05$ * |
| Time effect | $F(2,25)= 29.94$<br>$p < 0.001$ * | $F(2,25)= 18.37$<br>$p < 0.001$ * | $F(2,25)= 2.238$<br>$p = 0.128$ | $F(2,25)= 284,910$<br>$p < 0.001$ * | $F(2,25)= 0.219$<br>$p = 0.804$ |
| Exposure and time interaction | $F(2,25)= 81.57$<br>$p < 0.001$ * | $F(2,25)= 21.13$<br>$p < 0.001$ * | $F(2,25)= 23.26$<br>$p < 0.001$ * | $F(2,25)= 331,720$<br>$p < 0.001$ * | $F(2,25)= 1.268$<br>$p = 0.299$ |

**Table S2:** Results of two-way ANOVA statistical analyses for behavior parameters in mice treated with minocycline or clonazepam.

| | | Exposure and treatment factors – TWO-WAY ANOVA | | | Exposure to $CO_2$ – Student t-test |
|---|---|---|---|---|---|
| | | Exposure | Treatment | Interaction | Treatment |
| **Minocycline 40 mg.kg$^{-1}$** | **Jump** | F(1,60)= 42.216<br>p < 0.001 + | F(1,60)= 4.471<br>p = 0.039 * | F(1,60)= 4.471<br>p = 0.039 * | t(30)= 2.115<br>p = 0.042 * |
| | **Freezing** | F(1,60)= 301.787<br>p < 0.001 + | F(1,60)= 0.742<br>p = 0.3193 | F(1,60)= 0.742<br>p = 0.393 | t(30)= 0.861<br>p = 0.396 |
| | **Running** | F(1,60)= 87.463<br>p < 0.001 + | F(1,60)= 0.537<br>p = 0.466 | F(1,60)= 0.537<br>p = 0.466 | t(30)= 0.733<br>p = 0.469 |
| **Clonazepam 0.05 mg.kg$^{-1}$** | **Jump** | F(1,34)= 9.996<br>p = 0.003 + | F(1,34)= 8.663<br>p = 0.006 * | F(1,34)= 8.663<br>p = 0.006 * | t(17)= 2.938<br>p = 0.009 * |
| | **Freezing** | F(1,34)= 213.801<br>p < 0.001 + | F(1,34)= 8.624<br>p = 0.006 * | F(1,34)= 8.624<br>p = 0.006 * | t(17)= 2.937<br>p = 0.009 * |
| | **Running** | F(1,34)= 24.251<br>p < 0.001 + | F(1,34)= 9.554<br>p = 0.004 * | F(1,34)= 9.554<br>p = 0.004 * | t(17)= 3.091<br>p = 0.007 * |

+ normoxia ≠ $CO_2$ exposure
* Vehicle ≠ treatment group
### interaction between $CO_2$ exposure and treatment

**Table S3:** Results of two-way ANOVA statistical analyzes for ventilatory and metabolic parameters in mice treated with minocycline

**Minocycline 40 mg.kg$^{-1}$**

| **TWO-WAY ANOVA** | **Ventilatory and metabolic parameters – TWO-WAY ANOVA** | | | | | |
|---|---|---|---|---|---|---|
| | **$V_E$** | **$V_T$** | **fR** | **$VO_2$** | **$V_E/VO_2$** | **Tb** |
| Exposure effect | $F(1,58) = 206.023$<br>$p < 0.001$ * | $F(1,58) = 122.995$<br>$p < 0.001$ * | $F(1,58) = 16.126$<br>$p < 0.001$ * | $F(1,58) = 26.770$<br>$p < 0.001$ * | $F(1,58) = 83.736$<br>$p < 0.001$ * | $F(1,58) = 350.470$<br>$p < 0.001$ * |
| Treatment effect | $F(1,58) = 0.735$<br>$p = 0.395$ | $F(1,58) = 0.548$<br>$p = 0.462$ | $F(1,58) = 8.498$<br>$p = 0.005$ * | $F(1,58) = 0.012$<br>$p = 0.913$ | $F(1,58) = 2.009$<br>$p = 0.162$ | $F(1,58) = 0.198$<br>$p = 0.658$ |
| Exposure and treatment interaction | $F(1,58) = 0.119$<br>$p = 0.731$ | $F(1,58) = 0.232$<br>$p = 0.632$ | $F(1,58) = 1.535$<br>$p = 0.220$ | $F(1,58) = 2.676$<br>$p = 0.107$ | $F(1,58) = 3.669$<br>$p = 0.060$ | $F(1,58) = 3.909$<br>$p = 0.053$ * |

**Table S4:** Results of two-way ANOVA statistical analyzes for ventilatory and metabolic parameters in mice treated with clonazepam.

**Clonazepam 0.05 mg.kg$^{-1}$**

| TWO-WAY ANOVA | Ventilatory and metabolic parameters – TWO-WAY ANOVA | | | | | |
|---|---|---|---|---|---|---|
| | $V_E$ | $V_T$ | fR | $VO_2$ | $V_E/VO_2$ | Tb |
| Exposure effect | F(1,32)= 140.206<br>p < 0.001 * | F(1,32)= 108.861<br>p < 0.001 * | F(1,32)= 0.083<br>p < 0.001 * | F(1,32)= 373.429<br>p < 0.001 * | F(1,32)= 37.314<br>p < 0.001 * | F(1,32)= 347.153<br>p < 0.001 * |
| Treatment effect | F(1,32)= 0.708<br>p = 0.406 | F(1,32)= 0.534<br>p = 0.470 | F(1,32)= 3.176<br>p = 0.084 | F(1,32)= 0.890<br>p = 0.353 | F(1,32)= 0.270<br>p = 0.607 | F(1,32)= 0.025<br>p = 0.874 |
| Exposure and treatment interaction | F(1,32)= 0.072<br>p = 0.790 | F(1,32)= 1.499<br>p = 0.230 | F(1,32)= 1.026<br>p = 0.319 | F(1,32)= 1.273<br>p = 0.268 | F(1,32)= 0.001<br>p = 0.973 | F(1,32)= 0.095<br>p = 0.760 |

**Table S5:** Results of one-way ANOVA statistical analyses for cytokine analysis in mice treated with minocycline or clonazepam and exposure to normocapnia (A) and $CO_2$ (B).

**A) Cytokines analysis – Normocapnia – ONE-WAY ANOVA**

| **Cytokines plasma** | **IL-2sRα** | **IL-6** | **IL-10** | **TNFα** |
|---|---|---|---|---|
| Treatment | $F(2,26)= 0.422$<br>$p= 0.660$ | n.d | $F(2,27)= 15.279$<br>$p= 0.012$ * | n.d |
| **Cytokines LC** | **IL-2sRα** | **IL-6** | **IL-10** | **TNFα** |
| Treatment | $F(2,20)= 1.907$<br>$p = 0.175$ | $F(2,14)= 1.546$<br>$p = 0.247$ | $F(2,09)= 0.589$<br>$p = 0.575$ | $F(2,21)= 0.460$<br>$p = 0.637$ |

**B) Cytokines analysis – Exposure to $CO_2$ – ONE-WAY ANOVA**

| **Cytokines plasma** | **IL-2sRα** | **IL-6** | **IL-10** | **TNFα** |
|---|---|---|---|---|
| Treatment | $F(2,36)= 0.663$<br>$p= 0.522$ | $F(2,35)= 0.189$<br>$p= 0.828$ | $F(2,34)= 2.200$<br>$p= 0.126$ | n.d |
| **Cytokines LC** | **IL-2sRα** | **IL-6** | **IL-10** | **TNFα** |
| Treatment | $F(2,16)= 2.735$<br>$p = 0.095$ | $F(2,14)= 1.378$<br>$p = 0.284$ | $F(2,09)= 1.257$<br>$p = 0.330$ | $F(2,19)= 1.757$<br>$p = 0.199$ |

n.d: not detectable

* Vehicle ≠ clonazepam

**Table S6:** Results of two-way ANOVA statistical analyzes for cytokine analysis in mice treated with minocycline or clonazepam and exposured to $CO_2$.

**Cytokines analysis – Exposure and treatment factors – TWO-WAY ANOVA**

| | | Exposure | Treatment | Exposure and treatment interaction |
|---|---|---|---|---|
| **Cytokines plasma** | IL-2sRα | F(1,61)= 1.161<br>p= 0.286 | F(2,61)= 1.025<br>p= 0.365 | F(2,61)= 0.202<br>p= 0.817 |
| | IL-6 | F(1,62)= 13.450<br>p= 0.005 + | F(2,62)= 1.389<br>p= 0.257 | F(2,62)= 1.389<br>p= 0.257 |
| | IL-10 | F(1,61)= 0.339<br>p= 0.562 | F(2,61)= 3.717<br>p= 0.030 * | F(2,61)= 0.485<br>p= 0.618 |
| | TNFα | n.d | n.d | n.d |
| **Cytokines LC** | IL-2sRα | F(1,36)= 0.758<br>p = 0.389 | F(2,36)= 3.746<br>p = 0.033 # | F(2,36)= 0.213<br>p = 0.809 |
| | IL-6 | F(1,28)= 0.259<br>p = 0.614 | F(2,28)= 2.843<br>p = 0.075 | F(2,28)= 0.104<br>p = 0.901 |
| | IL-10 | F(1,18) = 0.337<br>p = 0.569 | F(2,18)= 0,172<br>p = 0.206 | F(2.18)= 0.500<br>p = 0.615 |
| | TNFα | F(1,41)= 2.195<br>p = 0.146 | F(2,41)= 1.650<br>p = 0.205 | F(2,41)= 0.894<br>p = 0.417 |

n.d: not detectable
+ normoxia Vehicle ≠ $CO_2$ exposure Vehicle
* $CO_2$ exposure Vehicle ≠ $CO_2$ exposure Minocycline
### normoxia Vehicle ≠ normoxia Clonazepam p= 0.088
